# Ethnic differences in receipt of psychological interventions in Early Intervention in Psychosis services in England: a cross-sectional study

**DOI:** 10.1101/2023.03.14.23287199

**Authors:** Merle Schlief, Nathalie Rich, Luke Sheridan Rains, Helen Baldwin, Antonio Rojas-Garcia, Patrick Nyikavaranda, Karen Persaud, Ceri Dare, Paul French, Brynmor Lloyd-Evans, Mike Crawford, Jo Smith, James B. Kirkbride, Sonia Johnson

## Abstract

**Background:** There is some evidence of inequitable psychosis care provision by ethnicity. We investigated variations in the receipt of CBTp and family intervention across ethnic groups in Early Intervention in Psychosis (EIP) teams throughout England, where national policy mandates offering these interventions to all.

**Methods:** We included data on 29,610 service users from the National Clinical Audit of Psychosis (NCAP), collected between 2018 and 2021. We conducted mixed effects logistic regression to examine odds ratios of receiving an intervention (CBTp, family intervention, or either intervention) across 17 ethnic groups while accounting for the effect of years and variance between teams and adjusting for individual- (age, gender, occupational status) and team-level covariates (care-coordinator caseload and mental health inequalities strategies).

**Findings:** Compared with White British people, every minoritized ethnic group, except those of mixed Asian-White and mixed Black African-White ethnicities, had lower adjusted odds of receiving CBTp (aOR 0·39, 95%CI 0·32-0·47 to 0·80, 0·64-1·00). People of Black African (0·61, 0·53-0·69), Black Caribbean (0·67, 0·56-0·81), non-African/Caribbean Black (0·63, 0·51-0·79), non-British/Irish White (0·73, 0·64-0·84), and of “any other” (0·66, 0·54-0·81) ethnicity also experienced lower adjusted odds of receiving family intervention.

**Interpretation:** Pervasive inequalities in receiving CBTp for first episode psychosis exist for almost all minoritized ethnic groups, and family intervention for many groups. Investigating how these inequalities arise should be a research priority, allowing co-produced development and testing of approaches to address them.

**Funding:** Independent research commissioned and funded by the National Institute for Health Research Policy Research Programme.

## Introduction

### Background

People from minoritized ethnic backgrounds often go through more complex and coercive pathways of psychosis care relative to their White counterparts.^1-5^ There is emerging evidence that these groups experience inequalities in the psychosis treatment they receive, including the use of pharmacological treatments and psychological interventions.^6-10^ People from minoritized ethnic backgrounds appear less likely to receive psychotherapy for psychosis, and to be referred for psychological treatments,^6-9^ which has been linked to increased likelihood of involuntary admissions.^11^ Which groups are affected and to what degree remains unclear, as do the stages of care at which inequalities emerge.

With a few exceptions,^12-14^ most studies from the US and UK observed ethnic inequalities in receipt of psychological interventions among people with psychosis, including first episode of psychosis (FEP).^6,7,15-19^ Studies from the UK found inequalities in offer and receipt of Cognitive Behavioural Therapy for psychosis (CBTp)^6,7,15,16,19^ and family intervention.^6^ While this was consistently the case for Black service users, evidence for other minoritized ethnic groups varied across studies including different samples and service settings.^6,7,16^ While most studies used audit data from one trust,^7,15,16,19^ a nationwide study using data from a National Audit of Schizophrenia (NAS) in England and Wales found all minoritized ethnic groups, except for those of mixed ethnicities, to be less likely to have been offered CBTp compared with White people.^6^ Black people were also less likely to have been offered family intervention, though Asian people were more likely.

A key setting for delivery of CBTp and family intervention are Early Intervention in Psychosis (EIP) services that have been available nationwide in England over the past 20 years. One policy-mandated goal is to offer all service users interventions that are recommended by National Institute for Health and Care Excellence (NICE) guidelines for people with psychosis.^20,21^ According to these guidelines, service users should receive 16 planned sessions of CBTp and 10 sessions of family intervention (given they are in close contact with their families), in conjunction with antipsychotic medication, as part of their individual treatment plan.^20,21^ A key question is whether ethnic inequalities are present in EIP services despite this commitment to an assertive and universal offer of interventions. No previous study has investigated differences in receipt of psychological interventions in EIP services for people with FEP specifically while also using fine-grained ethnic categories.

Substantial and high-quality evidence regarding ethnic differences in receipt of psychological and family intervention in EIP settings, where delivery is a policy requirement and service users are at a stage of symptoms that is important for long-term prognosis, is needed.

### Aim & Objectives

This study aimed to investigate the magnitude of inequalities in receipt of psychological interventions in EIP services.

Our specific objectives were:

1. To examine the association between ethnicity and receipt of CBTp, family intervention, and at least one of the two psychological interventions.
2. To investigate the role of other individual-level factors (age, gender, and occupational status), and team-level factors (whether teams have a strategy to address inequalities in mental health services and the average caseload of care coordinators) in the association between ethnicity and receipt of CBTp, family intervention, and either intervention.

## Methods

### Data collection

#### Participants

We used cross-sectional data from three years (2018/19 to 2020/21) of the National Clinical Audit of Psychosis (NCAP) commissioned across England by the Healthcare Quality Improvement Partnership (HQIP), a body working to promote quality across healthcare services.^22^ Data were collected retrospectively from all NHS EIP team via case-note audit of up to 100 randomly selected participants with FEP aged 14-65 years old, and a service-level questionnaire (*see* Supplement for further details on the NCAP methodology^23^). Each year, data were submitted for between 97% and 99% of the total expected number of EIP service users.^24-26^ Due to anonymization of NCAP audit data in each survey wave, it was not possible to identify participants who contributed to more than one wave. To be included in the audit, participants had to be on the caseload of an EIP team for at least six months at the census date. Participants were excluded from the audit if they experienced psychotic symptoms due to an organic cause or if they spent most of their time in a geographical area different from the EIP service.

#### Outcome

Our two main outcome variables were receipt of CBTp and family intervention. Receipt reflected both offer and uptake of appropriate and relevant care. We created a binary variable to indicate receipt of at least one session of each outcome in each year (*see* Supplement for further details). As a secondary outcome, we investigated receipt of either psychological intervention i.e. CBTp and/or family intervention.

#### Exposure and confounders

Our main exposure was ethnicity, grouped into 17 categories (as per NCAP methodology; *see* Table 1). Confounders included participant age (14-25, 26-35, 36-45, 46-55, 56-65) and gender (male, female, other), *a priori*, and participant occupational status (binary measure: in/out of work, education, or training at first assessment). We included two covariates at team-level, including whether the team or trust had a strategy to identify and address inequalities in mental health service use (yes/no), and a continuous proxy variable for staff caseload (mean number of service users per care coordinator).

**Table 1:**
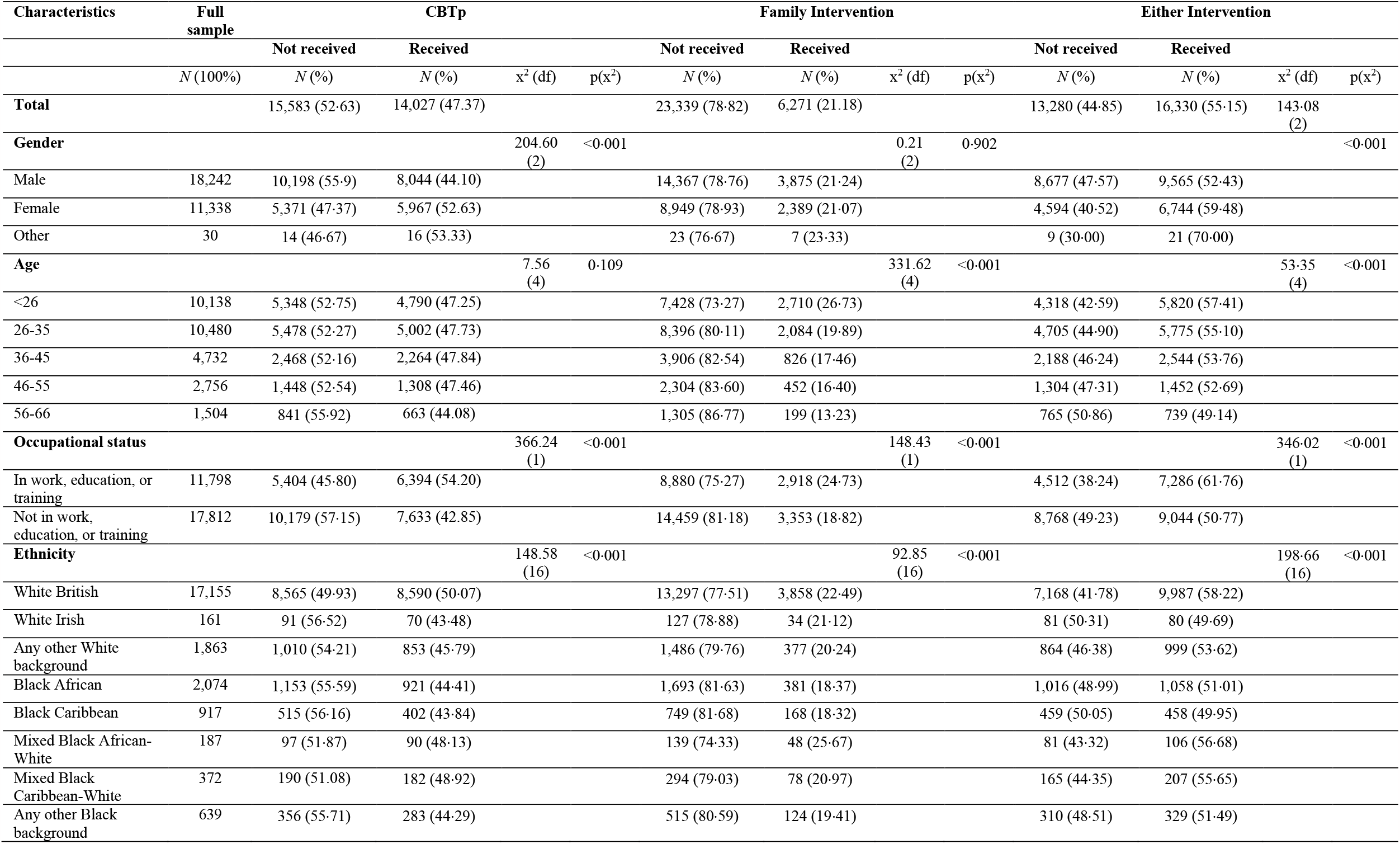
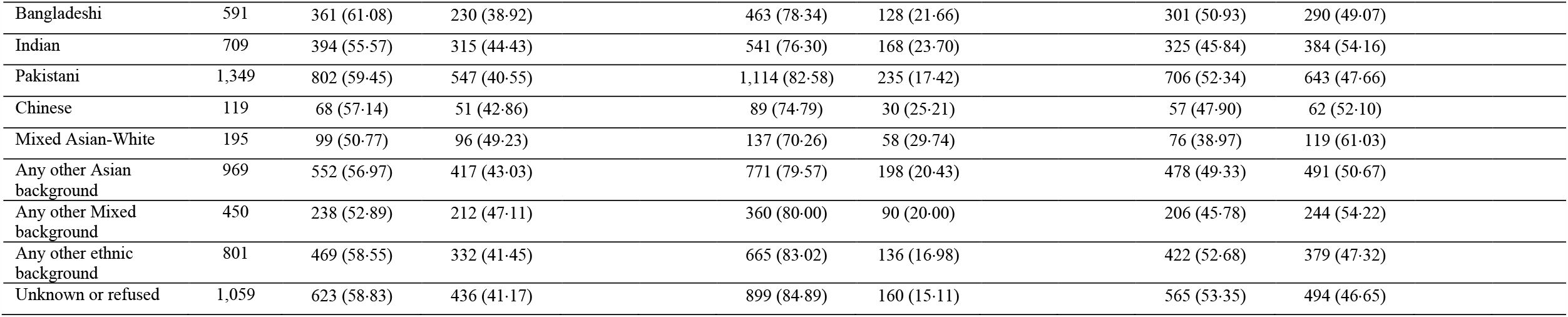
Sociodemographic characteristics and ethnicities, by receipt of CBTp, family intervention, and either intervention.

| Characteristics | Full sample | CBTp |  |  |  | Family Intervention |  |  |  | Either Intervention |  |  |  |
| --- | --- | --- | --- | --- | --- | --- | --- | --- | --- | --- | --- | --- | --- |
|  |  | Not received | Received |  |  | Not received | Received |  |  | Not received | Received |  |  |
| | *N* (100%) | *N* (%) | *N* (%) | $\chi^2$ (df) | *p*($\chi^2$) | *N* (%) | *N* (%) | $\chi^2$ (df) | *p*($\chi^2$) | *N* (%) | *N* (%) | $\chi^2$ (df) | *p*($\chi^2$) |
| **Total** |  | 15,583 (52.63) | 14,027 (47.37) |  |  | 23,339 (78.82) | 6,271 (21.18) |  |  | 13,280 (44.85) | 16,330 (55.15) | 143.08 (2) |  |
| **Gender** |  |  |  | 204.60 (2) | <0.001 |  |  | 0.21 (2) | 0.902 |  |  |  | <0.001 |
| Male | 18,242 | 10,198 (55.9) | 8,044 (44.10) |  |  | 14,367 (78.76) | 3,875 (21.24) |  |  | 8,677 (47.57) | 9,565 (52.43) |  |  |
| Female | 11,338 | 5,371 (47.37) | 5,967 (52.63) |  |  | 8,949 (78.93) | 2,389 (21.07) |  |  | 4,594 (40.52) | 6,744 (59.48) |  |  |
| Other | 30 | 14 (46.67) | 16 (53.33) |  |  | 23 (76.67) | 7 (23.33) |  |  | 9 (30.00) | 21 (70.00) |  |  |
| **Age** |  |  |  | 7.56 (4) | 0.109 |  |  | 331.62 (4) | <0.001 |  |  | 53.35 (4) | <0.001 |
| <26 | 10,138 | 5,348 (52.75) | 4,790 (47.25) |  |  | 7,428 (73.27) | 2,710 (26.73) |  |  | 4,318 (42.59) | 5,820 (57.41) |  |  |
| 26-35 | 10,480 | 5,478 (52.27) | 5,002 (47.73) |  |  | 8,396 (80.11) | 2,084 (19.89) |  |  | 4,705 (44.90) | 5,775 (55.10) |  |  |
| 36-45 | 4,732 | 2,468 (52.16) | 2,264 (47.84) |  |  | 3,906 (82.54) | 826 (17.46) |  |  | 2,188 (46.24) | 2,544 (53.76) |  |  |
| 46-55 | 2,756 | 1,448 (52.54) | 1,308 (47.46) |  |  | 2,304 (83.60) | 452 (16.40) |  |  | 1,304 (47.31) | 1,452 (52.69) |  |  |
| 56-66 | 1,504 | 841 (55.92) | 663 (44.08) |  |  | 1,305 (86.77) | 199 (13.23) |  |  | 765 (50.86) | 739 (49.14) |  |  |
| **Occupational status** |  |  |  | 366.24 (1) | <0.001 |  |  | 148.43 (1) | <0.001 |  |  | 346.02 (1) | <0.001 |
| In work, education, or training | 11,798 | 5,404 (45.80) | 6,394 (54.20) |  |  | 8,880 (75.27) | 2,918 (24.73) |  |  | 4,512 (38.24) | 7,286 (61.76) |  |  |
| Not in work, education, or training | 17,812 | 10,179 (57.15) | 7,633 (42.85) |  |  | 14,459 (81.18) | 3,353 (18.82) |  |  | 8,768 (49.23) | 9,044 (50.77) |  |  |
| **Ethnicity** |  |  |  | 148.58 (16) | <0.001 |  |  | 92.85 (16) | <0.001 |  |  | 198.66 (16) | <0.001 |
| White British | 17,155 | 8,565 (49.93) | 8,590 (50.07) |  |  | 13,297 (77.51) | 3,858 (22.49) |  |  | 7,168 (41.78) | 9,987 (58.22) |  |  |
| White Irish | 161 | 91 (56.52) | 70 (43.48) |  |  | 127 (78.88) | 34 (21.12) |  |  | 81 (50.31) | 80 (49.69) |  |  |
| Any other White background | 1,863 | 1,010 (54.21) | 853 (45.79) |  |  | 1,486 (79.76) | 377 (20.24) |  |  | 864 (46.38) | 999 (53.62) |  |  |
| Black African | 2,074 | 1,153 (55.59) | 921 (44.41) |  |  | 1,693 (81.63) | 381 (18.37) |  |  | 1,016 (48.99) | 1,058 (51.01) |  |  |
| Black Caribbean | 917 | 515 (56.16) | 402 (43.84) |  |  | 749 (81.68) | 168 (18.32) |  |  | 459 (50.05) | 458 (49.95) |  |  |
| Mixed Black African-White | 187 | 97 (51.87) | 90 (48.13) |  |  | 139 (74.33) | 48 (25.67) |  |  | 81 (43.32) | 106 (56.68) |  |  |
| Mixed Black Caribbean-White | 372 | 190 (51.08) | 182 (48.92) |  |  | 294 (79.03) | 78 (20.97) |  |  | 165 (44.35) | 207 (55.65) |  |  |
| Any other Black background | 639 | 356 (55.71) | 283 (44.29) |  |  | 515 (80.59) | 124 (19.41) |  |  | 310 (48.51) | 329 (51.49) |  |  |

|  |  |  |  |  |  |  |  |
| --- | --- | --- | --- | --- | --- | --- | --- |
| Bangladeshi | 591 | 361 (61.08) | 230 (38.92) | 463 (78.34) | 128 (21.66) | 301 (50.93) | 290 (49.07) |
| Indian | 709 | 394 (55.57) | 315 (44.43) | 541 (76.30) | 168 (23.70) | 325 (45.84) | 384 (54.16) |
| Pakistani | 1,349 | 802 (59.45) | 547 (40.55) | 1,114 (82.58) | 235 (17.42) | 706 (52.34) | 643 (47.66) |
| Chinese | 119 | 68 (57.14) | 51 (42.86) | 89 (74.79) | 30 (25.21) | 57 (47.90) | 62 (52.10) |
| Mixed Asian-White | 195 | 99 (50.77) | 96 (49.23) | 137 (70.26) | 58 (29.74) | 76 (38.97) | 119 (61.03) |
| Any other Asian background | 969 | 552 (56.97) | 417 (43.03) | 771 (79.57) | 198 (20.43) | 478 (49.33) | 491 (50.67) |
| Any other Mixed background | 450 | 238 (52.89) | 212 (47.11) | 360 (80.00) | 90 (20.00) | 206 (45.78) | 244 (54.22) |
| Any other ethnic background | 801 | 469 (58.55) | 332 (41.45) | 665 (83.02) | 136 (16.98) | 422 (52.68) | 379 (47.32) |
| Unknown or refused | 1,059 | 623 (58.83) | 436 (41.17) | 899 (84.89) | 160 (15.11) | 565 (53.35) | 494 (46.65) |

### Statistical Analysis

We conducted a complete case analysis, excluding participants with missing data. We compared differences between these two groups using *Chi*-squared or Fisher’s exact tests, and similarly reported descriptive statistics on the complete case sample, and any differences by outcome status. Next, we used multilevel logistic regression models to examine the association between each outcome and ethnicity, adjusted for confounders. We included a random intercept for EIP team to account for potential team-level variation in treatment, and random slopes between EIP team and survey year to account for any yearly differences in variance of psychological interventions attributable to the EIP team level, e.g., due to COVID-19. For each outcome, we reported findings from null, age-sex adjusted, and fully adjusted models, including odds ratios (OR) and 95% confidence intervals (95%CI) for ethnic variation in receipt of psychological interventions and intraclass correlation coefficients (ICC) of the proportion of outcomes attributable to the team-level (*see* Supplement for further details).

We conducted sensitivity analyses re-running our main model for each primary outcome in separate survey years to account for potential sample overlap. We also conducted sensitivity analyses to investigate ethnic variation in “offer” rather than “receipt” of psychological interventions (only available in 2019/20 and 2020/21 survey years). Here, we fitted our final multivariable models from our primary analyses of “receipt” for “offer” outcomes.

## Results

The final analytic sample included 29,610 participants (Figure S1), after excluding 1·69% (n=510). Excluded participants did not differ from the complete case sample by outcome status, gender, age, or occupational status, but did differ by ethnicity (p<0·001), with people from a Black Caribbean (6·33%) and “any other Black” background (5·47%) having the highest percentage of missingness, and people from of White British (0·64%) and Chinese (0·83%) ethnicity having the lowest percentage of missingness (Table S1).

### Descriptive statistics

The majority of the sample was White British (57·94%), male (61·61%), aged 35 or younger (69·63%), and not in work, education, or training (60·16%) (Table 1). The overall proportion of people who received CBTp and family interventions were 47·37% and 21·18%, respectively. Receipt of CBTp differed by gender, occupational status, and ethnicity, and receipt of family intervention by age, occupational status, and ethnicity (*see* Table 1 and Supplement for further details). Compared with receipt, the overall proportion of the sample offered CBTp and family interventions were higher, at 80·97% and 62·27%, respectively (Table S2).

### Multilevel modelling

#### Ethnic variation in receipt of CBTp

In a null model, 18·53% of the variance in receipt of CBTp was attributable to the EIP team level (95%CI: 0·14-0·23), when holding the random slope constant, and increased slightly in a model adjusted for ethnicity and other fixed effects covariates (ICC 0·21; 95%CI: 0·16-0·26). Unadjusted and adjusted results were similar with respect to ethnicity (Table 2). In the adjusted model, people of Bangladeshi ethnicity had the lowest odds of receiving CBTp (aOR: 0·39; 95%CI: 0·32-0·47), followed by those of Chinese (aOR: 0·52; 95%CI: 0·35-0·77), Black African (aOR: 0·53; 95%CI: 0·47-0·59), and Pakistani (aOR: 0·54; 95%CI: 0·48-0·62) ethnicity relative to White British participants. Participants who were female (aOR: 1·42; 95%CI: 1·35-1·50) and in work, education or training (aOR: 1·63; 95%CI: 1·55-1·72) were more likely to receive CBTp, while those below 26-years-old (aOR: 0·91; 95%CI: 0·86-0·97) or above 46-years-old were less likely to receive CBTp compared with those aged 26-35-years-old (Table 2). Finally, those in teams with an inequalities strategy (aOR: 1·19; 95%CI: 1·04-1·37) had higher odds of receiving CBTp, but caseload size was not associated with receipt of CBTp (per one extra patient per care coordinate: aOR: 0·99; 95%CI: 0·97-1·00).

**Table 2:**
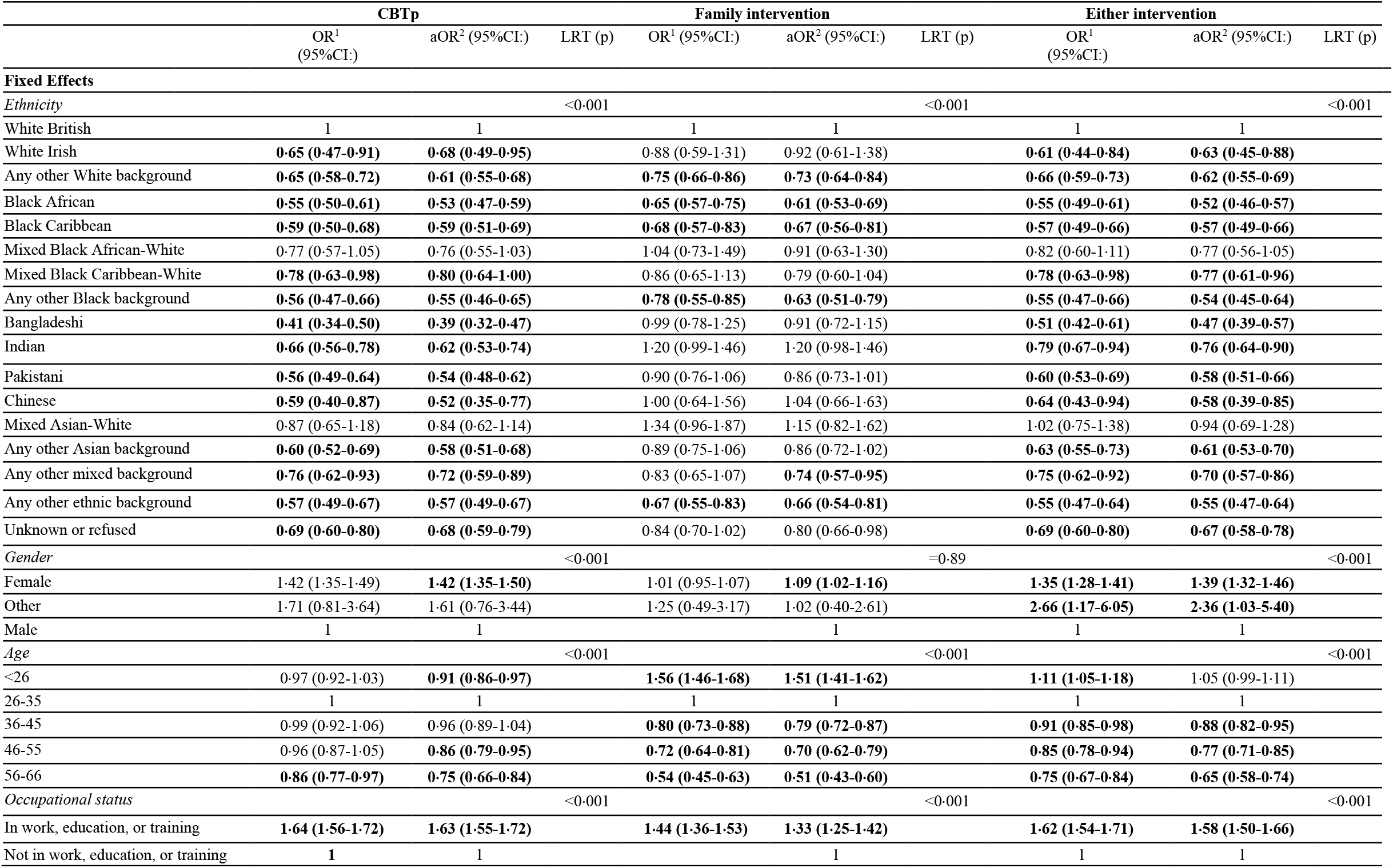
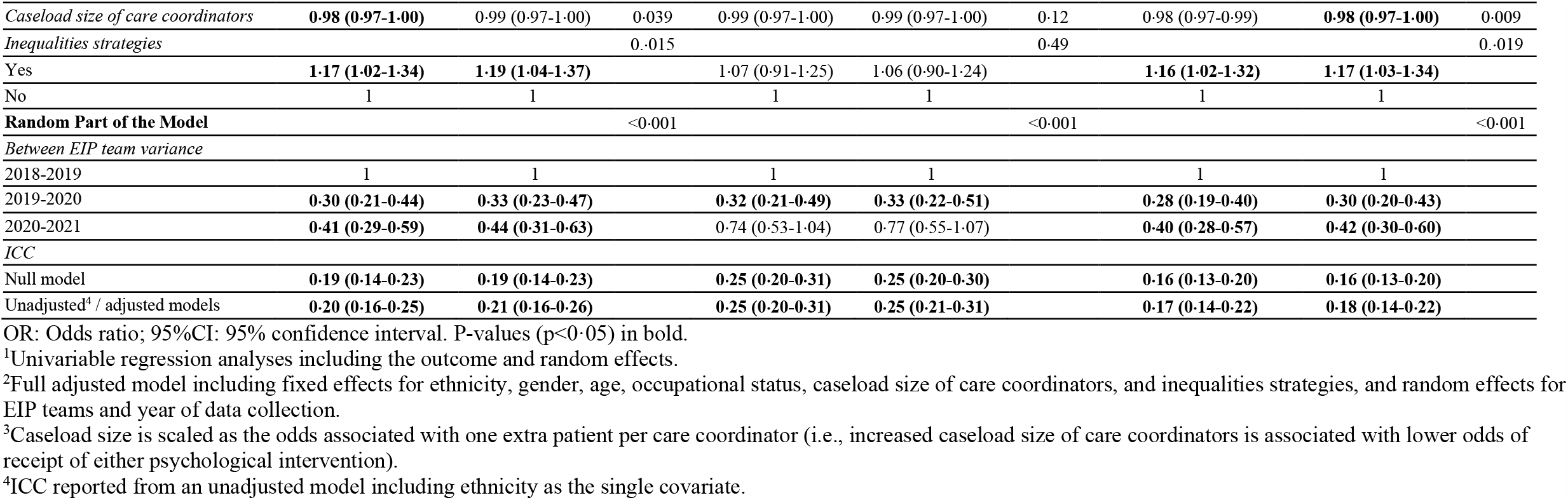
Association between ethnicities and receipt of CBTp, family intervention, and either intervention.

|  | CBTp |  |  | Family intervention |  |  | Either intervention |  |  |
| --- | --- | --- | --- | --- | --- | --- | --- | --- | --- |
|  | OR<sup>1</sup><br>(95%CI:) | aOR<sup>2</sup> (95%CI:) | LRT (p) | OR<sup>1</sup> (95%CI:) | aOR<sup>2</sup> (95%CI:) | LRT (p) | OR<sup>1</sup><br>(95%CI:) | aOR<sup>2</sup> (95%CI:) | LRT (p) |
| **Fixed Effects** |  |  |  |  |  |  |  |  |  |
| *Ethnicity* |  |  | <0.001 |  |  | <0.001 |  |  | <0.001 |
| White British | 1 | 1 |  | 1 | 1 |  | 1 | 1 |  |
| White Irish | **0.65 (0.47-0.91)** | **0.68 (0.49-0.95)** |  | 0.88 (0.59-1.31) | 0.92 (0.61-1.38) |  | **0.61 (0.44-0.84)** | **0.63 (0.45-0.88)** |  |
| Any other White background | **0.65 (0.58-0.72)** | **0.61 (0.55-0.68)** |  | **0.75 (0.66-0.86)** | **0.73 (0.64-0.84)** |  | **0.66 (0.59-0.73)** | **0.62 (0.55-0.69)** |  |
| Black African | **0.55 (0.50-0.61)** | **0.53 (0.47-0.59)** |  | **0.65 (0.57-0.75)** | **0.61 (0.53-0.69)** |  | **0.55 (0.49-0.61)** | **0.52 (0.46-0.57)** |  |
| Black Caribbean | **0.59 (0.50-0.68)** | **0.59 (0.51-0.69)** |  | **0.68 (0.57-0.83)** | **0.67 (0.56-0.81)** |  | **0.57 (0.49-0.66)** | **0.57 (0.49-0.66)** |  |
| Mixed Black African-White | 0.77 (0.57-1.05) | 0.76 (0.55-1.03) |  | 1.04 (0.73-1.49) | 0.91 (0.63-1.30) |  | 0.82 (0.60-1.11) | 0.77 (0.56-1.05) |  |
| Mixed Black Caribbean-White | **0.78 (0.63-0.98)** | **0.80 (0.64-1.00)** |  | 0.86 (0.65-1.13) | 0.79 (0.60-1.04) |  | **0.78 (0.63-0.98)** | **0.77 (0.61-0.96)** |  |
| Any other Black background | **0.56 (0.47-0.66)** | **0.55 (0.46-0.65)** |  | **0.78 (0.55-0.85)** | **0.63 (0.51-0.79)** |  | **0.55 (0.47-0.66)** | **0.54 (0.45-0.64)** |  |
| Bangladeshi | **0.41 (0.34-0.50)** | **0.39 (0.32-0.47)** |  | 0.99 (0.78-1.25) | 0.91 (0.72-1.15) |  | **0.51 (0.42-0.61)** | **0.47 (0.39-0.57)** |  |
| Indian | **0.66 (0.56-0.78)** | **0.62 (0.53-0.74)** |  | 1.20 (0.99-1.46) | 1.20 (0.98-1.46) |  | **0.79 (0.67-0.94)** | **0.76 (0.64-0.90)** |  |
| Pakistani | **0.56 (0.49-0.64)** | **0.54 (0.48-0.62)** |  | 0.90 (0.76-1.06) | 0.86 (0.73-1.01) |  | **0.60 (0.53-0.69)** | **0.58 (0.51-0.66)** |  |
| Chinese | **0.59 (0.40-0.87)** | **0.52 (0.35-0.77)** |  | 1.00 (0.64-1.56) | 1.04 (0.66-1.63) |  | **0.64 (0.43-0.94)** | **0.58 (0.39-0.85)** |  |
| Mixed Asian-White | 0.87 (0.65-1.18) | 0.84 (0.62-1.14) |  | 1.34 (0.96-1.87) | 1.15 (0.82-1.62) |  | 1.02 (0.75-1.38) | 0.94 (0.69-1.28) |  |
| Any other Asian background | **0.60 (0.52-0.69)** | **0.58 (0.51-0.68)** |  | 0.89 (0.75-1.06) | 0.86 (0.72-1.02) |  | **0.63 (0.55-0.73)** | **0.61 (0.53-0.70)** |  |
| Any other mixed background | **0.76 (0.62-0.93)** | **0.72 (0.59-0.89)** |  | 0.83 (0.65-1.07) | **0.74 (0.57-0.95)** |  | **0.75 (0.62-0.92)** | **0.70 (0.57-0.86)** |  |
| Any other ethnic background | **0.57 (0.49-0.67)** | **0.57 (0.49-0.67)** |  | **0.67 (0.55-0.83)** | **0.66 (0.54-0.81)** |  | **0.55 (0.47-0.64)** | **0.55 (0.47-0.64)** |  |
| Unknown or refused | **0.69 (0.60-0.80)** | **0.68 (0.59-0.79)** |  | 0.84 (0.70-1.02) | 0.80 (0.66-0.98) |  | **0.69 (0.60-0.80)** | **0.67 (0.58-0.78)** |  |
| *Gender* |  |  | <0.001 |  |  | =0.89 |  |  | <0.001 |
| Female | 1.42 (1.35-1.49) | **1.42 (1.35-1.50)** |  | 1.01 (0.95-1.07) | **1.09 (1.02-1.16)** |  | **1.35 (1.28-1.41)** | **1.39 (1.32-1.46)** |  |
| Other | 1.71 (0.81-3.64) | 1.61 (0.76-3.44) |  | 1.25 (0.49-3.17) | 1.02 (0.40-2.61) |  | **2.66 (1.17-6.05)** | **2.36 (1.03-5.40)** |  |
| Male | 1 | 1 |  | 1 | 1 |  | 1 | 1 |  |
| *Age* |  |  | <0.001 |  |  | <0.001 |  |  | <0.001 |
| <26 | 0.97 (0.92-1.03) | **0.91 (0.86-0.97)** |  | **1.56 (1.46-1.68)** | **1.51 (1.41-1.62)** |  | **1.11 (1.05-1.18)** | 1.05 (0.99-1.11) |  |
| 26-35 | 1 | 1 |  | 1 | 1 |  | 1 | 1 |  |
| 36-45 | 0.99 (0.92-1.06) | 0.96 (0.89-1.04) |  | **0.80 (0.73-0.88)** | **0.79 (0.72-0.87)** |  | **0.91 (0.85-0.98)** | **0.88 (0.82-0.95)** |  |
| 46-55 | 0.96 (0.87-1.05) | **0.86 (0.79-0.95)** |  | **0.72 (0.64-0.81)** | **0.70 (0.62-0.79)** |  | **0.85 (0.78-0.94)** | **0.77 (0.71-0.85)** |  |
| 56-66 | **0.86 (0.77-0.97)** | **0.75 (0.66-0.84)** |  | **0.54 (0.45-0.63)** | **0.51 (0.43-0.60)** |  | **0.75 (0.67-0.84)** | **0.65 (0.58-0.74)** |  |
| *Occupational status* |  |  | <0.001 |  |  | <0.001 |  |  | <0.001 |
| In work, education, or training | **1.64 (1.56-1.72)** | **1.63 (1.55-1.72)** |  | **1.44 (1.36-1.53)** | **1.33 (1.25-1.42)** |  | **1.62 (1.54-1.71)** | **1.58 (1.50-1.66)** |  |
| Not in work, education, or training | 1 | 1 |  | 1 | 1 |  | 1 | 1 |  |
| *Caseload size of care coordinators* | **0.98 (0.97-1.00)** | 0.99 (0.97-1.00) | 0.039 | 0.99 (0.97-1.00) | 0.99 (0.97-1.00) | 0.12 | 0.98 (0.97-0.99) | **0.98 (0.97-1.00)** | 0.009 |
| *Inequalities strategies* |  |  | 0.015 |  |  | 0.49 |  |  | 0.019 |
| Yes | **1.17 (1.02-1.34)** | **1.19 (1.04-1.37)** |  | 1.07 (0.91-1.25) | 1.06 (0.90-1.24) |  | **1.16 (1.02-1.32)** | **1.17 (1.03-1.34)** |  |
| No | 1 | 1 |  | 1 | 1 |  | 1 | 1 |  |
| **Random Part of the Model** |  |  | <0.001 |  |  | <0.001 |  |  | <0.001 |
| *Between EIP team variance* |  |  |  |  |  |  |  |  |  |
| 2018-2019 | 1 | 1 |  | 1 | 1 |  | 1 | 1 |  |
| 2019-2020 | **0.30 (0.21-0.44)** | **0.33 (0.23-0.47)** |  | **0.32 (0.21-0.49)** | **0.33 (0.22-0.51)** |  | **0.28 (0.19-0.40)** | **0.30 (0.20-0.43)** |  |
| 2020-2021 | **0.41 (0.29-0.59)** | **0.44 (0.31-0.63)** |  | 0.74 (0.53-1.04) | 0.77 (0.55-1.07) |  | **0.40 (0.28-0.57)** | **0.42 (0.30-0.60)** |  |
| *ICC* |  |  |  |  |  |  |  |  |  |
| Null model | **0.19 (0.14-0.23)** | **0.19 (0.14-0.23)** |  | **0.25 (0.20-0.31)** | **0.25 (0.20-0.30)** |  | **0.16 (0.13-0.20)** | **0.16 (0.13-0.20)** |  |
| Unadjusted<sup>4</sup> / adjusted models | **0.20 (0.16-0.25)** | **0.21 (0.16-0.26)** |  | **0.25 (0.20-0.31)** | **0.25 (0.21-0.31)** |  | **0.17 (0.14-0.22)** | **0.18 (0.14-0.22)** |  |

OR: Odds ratio; 95%CI: 95% confidence interval. P-values (p<0.05) in bold.
<sup>1</sup>Univariable regression analyses including the outcome and random effects.
<sup>2</sup>Full adjusted model including fixed effects for ethnicity, gender, age, occupational status, caseload size of care coordinators, and inequalities strategies, and random effects for EIP teams and year of data collection.
<sup>3</sup>Caseload size is scaled as the odds associated with one extra patient per care coordinator (i.e., increased caseload size of care coordinators is associated with lower odds of receipt of either psychological intervention).
<sup>4</sup>ICC reported from an unadjusted model including ethnicity as the single covariate.

#### Ethnic variation in receipt of family interventions

Compared with CBTp, a greater portion of variance in receipt of family intervention was attributable to the EIP team level in the null model (ICC 0·25; 95%CI: 0·20-0·31), which persisted in unadjusted and adjusted models (Table 2). Following multivariable adjustment models, six minoritized ethnic groups were less likely to receive family intervention (Table 2), including those of Black African (aOR: 0·61; 95%CI: 0·53-0·69), Black Caribbean (aOR: 0·67; 95%CI: 0·56-0·81), “any other Black” (aOR: 0·63; 95%CI: 0·51-0·79), “any other White” (aOR: 0·73; 95%CI: 0·64-0·84), “any other mixed” (aOR: 0·74; 95%CI: 0·57-0·95), or “any other” (aOR: 0·66; 95%CI: 0·54-0·81) ethnicities. Being female (aOR: 1·09; 95%CI: 1·02-1·16), not in work, education, or training (aOR: 1·33; 95%CI: 1·25-1·42), or younger than 26 years old (aOR: 1·51; 95%CI: 1·41-1·62) was associated with higher odds, and being older than 35-years-old with lower odds of receiving family intervention (Table 2). We did not observe differences in receipt of family intervention by average caseload size or presence of an inequalities strategy.

#### Ethnic variation in receipt of either psychological intervention

Results from this model were similar to CBTp, with additional weak evidence that participants in EIP teams with greater caseloads were less likely to have received either intervention (aOR: 0·98; 95%CI: 0·97-1·00; Table 2).

#### Sensitivity analyses

##### Receipt of CBTp, family intervention, and either intervention using separate samples by year

In the sensitivity analyses examining each year of data collection separately, odds of receipt of CBTp were marginally larger for some ethnic groups and smaller for others compared with those of the merged sample (Tables S3-5). Differences in receipt of CBTp failed to reach significance for some ethnic groups, including White Irish across all three years, mixed Black Caribbean-White in 2018-2019 and 2020-2021, and Chinese and “any other mixed” ethnicities in 2019-2020, but were larger in the other years for the latter two groups.

Odds of receipt of family intervention marginally differed in both directions across years compared to the merged sample (Tables S3-5). Odds failed to reach significance for the Black Caribbean group in 2018-2019, “any other” ethnic background in 2018-2019 and 2019-2020, and “White other” and “any other mixed” background in 2018-2020 and 2020-2021 but were marginally larger compared to the main sample in other years. Contrary to the merged sample, odds reached significance for the mixed Black Caribbean-White group in 2019-2020 and “any other Asian” background in 2020-2021.

Odds of receipt of either intervention failed to reach significance for the Indian group in 2018-2019 and 2019-2020, White Irish group in 2018-2019 and 2019-2020, mixed Black Caribbean-White group in 2018-2019 and 2020-2021, and “any other mixed” background in 2020-2021 (Tables S3-5). However, differences in receipt of either intervention were larger in the other years for these ethnic groups compared to the merged sample. Odds were not significant for the Chinese group across years.

##### Offer of CBTp, family intervention, and either intervention using the merged sample

Patterns of ethnic disparities in *offer* of CBTp (Table S6) were broadly similar to those for *receipt* (Table 2), with point estimates tending to indicate less reductions in *offer* (*see* Supplement for further details). Exceptions to this existed, and (unlike receipt) we observed no statistically significant differences in offer of CBTp between White British and Black Caribbean, mixed Black Caribbean-White, and Chinese participants. Odds reached significance for people from a mixed Asian-White background (aOR: 0·51; 95%CI: 0·51-0·82).

Patterns of ethnic disparities in *offer* of family interventions (Table S6) were broadly similar to those for *receipt* (Table 2). Exceptions existed, and (unlike receipt) we observed no statistically significant differences in offer of family interventions for those of “any other Black” background, and found that those from an Indian background were more likely to be offered family intervention compared with White British participants (aOR: 1.26; 95%CI: 1·01-1·56).

Patterns of ethnic difference in *offer* of either intervention (Table S6) resembled those of *receipt* for most ethnic groups (Table 2). However, contrary to receipt, we observed no statistically significant differences in *offer* of either intervention for six ethnic groups, and significantly lower odds among those of mixed Asian-White background (aOR: 0·50; 95%CI: 0·29-0·87).

## Discussion

This is the first cross-sectional study to examine differences in receipt of psychological interventions among people with FEP of several specific ethnicities using nationwide data from EIP services mandated to deliver these interventions to all service users. We found evidence for inequalities in receipt of psychological interventions, including both CBTp and family intervention independently, across most ethnic groups. This was most consistent for CBTp, where the odds of receiving CBTp were reduced by between 20-61% for most minoritized ethnic groups, after adjustment for covariates. Decreased odds were most pronounced for Bangladeshi groups, but Pakistani, Chinese, and Black African patients were also almost half as likely to receive CBTp as their White British counterparts. Receipt of family intervention were notably reduced by between 33-39% in Black Caribbean, Black African, and other Black groups. Other differences in receipt of psychological interventions included reduced odds of receipt amongst those older than 35/45-years-old, and greater odds of receipt for women and service users who were in work, education, or training, as well as amongst those in EIP teams who reported having an inequalities strategy in place. Variance at team-level accounted for approximately 19-25% in differences in receipt of interventions and remained similar when controlling for relevant covariates. When examining differences in which service users were reported to have been offered, as opposed to receiving, CBTp or family intervention, patterns were largely similar. Compared to receipt, differences in offer of interventions were marginally smaller for most ethnic groups and failed to reach significance for a few.

### Limitations

The study’s findings should be considered in the context of following methodological limitations. First, we were unable to control for potential sample overlap between the three audit years which were merged for our main analyses, potentially inflating observed effect sizes. When the three years of data are analysed separately, odds ratios for some ethnic groups varied in size and failed to reach significance compared to the merged sample. However, findings remained comparable for most ethnic groups. Second, we conducted a complete case analysis excluding anyone with missing data on the exposure, outcomes, or covariates. We expect the complete case analysis to produce unbiased results based on the low percentage of excluded cases (1·69% of all participants).^27^ Third, while we explored the role of several individual-level and team-level covariates, we did not account for variance between NHS trusts, and were limited to covariates available in the NCAP audit dataset. We were not able to include other potentially relevant covariates, including area-level deprivation, which might partially account for the observed differences in receipt of psychological interventions. Lastly, outcomes were based on clinician rather than service user reports potentially impacting the reliability of reporting, especially regarding the offer of interventions.

### Findings in context of previous studies

#### Ethnic inequalities

##### CBTp

We found evidence for lower odds of receiving CBTp, and also of being offered CBTp, among most minoritized ethnic groups, except of mixed Black African-White and mixed Asian-White ethnicity, and obtained similar results for offer of CBTp even though odds ratios marginally increased for most ethnic groups and failed to reach significance for Black Caribbean and mixed Black Caribbean-White groups. This is in line with previous UK and most US studies which have found lower odds of being offered or receiving CBTp or other psychotherapy among service users of Black ethnicity.^6,7,15-19^ Findings for other ethnic groups have been more mixed.^6,7,16-18^ In accordance with our findings, most studies, including a UK study using nationwide audit data, also showed differences in care for other minoritized ethnic groups.^6,16,18^ Divergences in findings across studies may reflect differences in samples, services, and covariates examined, including potential variations in factors associated with care, such as diversity of staff and aspects of service delivery intended to improve noted inequalities. Overall, findings confirm significant inequalities in both being offered and receiving psychological interventions, and indicate that these are present at an early stage in treatment, and despite a national policy requirement to deliver equitable EIP services to all.

### Family intervention

In line with the findings of a study using nationwide audit data, we found lower odds of receiving and being offered family intervention among people from Black Caribbean and Black African, compared to White British people, but no differences among most mixed ethnic groups.^6^ Service users from “any other Black” ethnic background had lower odds of receipt only. Das-Munshi and colleagues also found higher odds of being offered family intervention among Asian and Asian British compared to White British people.^6^ We did not find evidence for differences in receipt of family intervention between people of different Asian ethnic backgrounds and White British people overall, however, people of Indian ethnicity had greater odds of being offered family intervention. Deviations in findings might be due to differences in service settings and ethnic categories explored with our study looking at EIP services only and more fine-grained, but smaller ethnic groups. Our findings further suggested that White people other than White Irish or White British people had lower odds of receiving and being offered family intervention. Lower odds of receiving and being offered family intervention may also reflect a lower likelihood of living or being in closely in contact with families early in the course of psychosis among certain ethnic groups.

### Potential explanations for inequalities in receipt of care

#### Ethnic inequalities

Ethnic inequalities in psychological interventions may arise at several points in the care pathway. We found evidence for inequalities in both receipt and offer of psychological interventions. While differences in offer of interventions were (marginally) smaller than differences in receipt for most ethnic groups, this suggests staff factors and service structures play an important role in inequalities in care. Whether an intervention is offered may reflect clinicians’ (mis)perceptions of appropriateness of care for different groups,^28^ an institutionally racist culture, and/or service capacity limitations, including availability of interpreters.^28-30^ Minoritized ethnic groups, specifically Black ethnic groups, have been found to be subject to more coercive pathways to care and present as more severe at first diagnosis in the UK.^4,5,31^ As a result, the window for psychological intervention offer might be missed by some minoritized ethnic groups due to being perceived as too unwell to benefit from psychological interventions or where psychological interventions may not be prioritised for compulsorily admitted patients in hospital wards compared to antipsychotic medication, despite NICE guidelines recommending psychological interventions during the acute phase of symptoms.^21^ Additionally, people from minoritized backgrounds may be less likely to engage with care due to negative experiences of services restricting opportunities to offer therapy interventions. Interestingly, whether a team had a strategy to address mental health inequalities in place was associated with receipt of CBTp, but not offer. Thus, existence of a strategy might influence service factors related to uptake rather than impacting offer of interventions among clinicians.

Whether service users accept an offer of treatment may be influenced by attitudes towards mental health problems and psychological treatments, and this may also influence whether they remain engaged with the services. This may be influenced by cultural differences in beliefs and stigma^28-30,32^ as well as individual and community experiences of services, including coercive pathways to care,^29,33^ and cultural ignorance and racism among clinicians,^28,29,32,34^ leading to mistrust towards professionals and services.^28,29,33^ Uptake may further reflect clinicians’ ability to offer and explain treatments in a way that appears acceptable and relevant to people from a range of backgrounds, as may the quality and cultural appropriateness of informational materials about treatments, including whether they have been adapted and co-produced with people from the relevant background.^28,29^ The appropriateness and acceptability of family intervention is also likely to be influenced by family composition, language proficiency, and availability, and by attitudes among family members and communities.^30^

#### Age and occupational inequalities

Our study found evidence for inequalities in care across different age groups. Lower odds of receiving family intervention or CBTp among age groups 36 or 46 and older may be explained by the only recent expansion of EIP services in England to older age groups (36-65 years) recommended by NICE in 2016.^35^ Previous studies showed that older EIP service users differed regarding service use needs, referral route, and duration of untreated psychosis potentially accounting for differences in receipt of care and indicating a lack of tailored interventions provided to older age groups.^36-38^ Our study also found greater odds of receiving family intervention among people younger than 26. Young people might live at home or near their parents, facilitating their involvement in care. Lastly, we found lower odds of receiving psychological interventions among people who were not in work, education, or training compared to those who were. Service users who are not in occupation at first presentation may experience more severe symptoms, lower general functioning, and longer duration of untreated psychosis.^39^ This might hinder engagement in work/education and care,^40^ and they might be perceived as being less able to accept interventions.

### Implications for research and practice

Our results highlight that, at a national level, most minoritized ethnic groups are offered and receive psychological interventions in EIP services less often than White British people. Co-produced, qualitative studies including staff and service users are needed to shed light on the underlying reasons for inequalities in care across different ethnic group and on approaches to addressing these. The finding of marked differences in offers and receipt of therapies makes addressing the question of why people from minoritized backgrounds are less likely to be offered therapy especially pressing. Further investigation is also indicated to understand differences between teams, including area-level factors such as deprivation, ethnic density, and urbanicity, and service factors, such as ethnic diversity among service users and staff of teams, or leadership and organisational context in teams.

Mandating inequalities strategies as part of routine EIP care might reduce ethnic inequalities in care. However, we found receipt, but not offer, of CBTp and either intervention to be higher in EIP teams which reported to have an inequalities strategy in place. Thus, existence of strategies might be an indicator of ethnic diversity of cases and service resources, rather than impacting on offer of care. More research is needed on the type and implementation of strategies addressing inequalities in receipt and offer of care. For instance, South London and Maudsely (SLaM) trust has a Patient and Carer Race Equality Framework (PCREF) taskforce, which was formed to address longstanding ethnic inequalities in care.^41,42^

Lastly, more evidence is needed on inequalities in other NICE mandated treatments, including education and employment support, physical health monitoring and interventions, carer education and support, and prescribing of antipsychotic medication with lack of UK nation-wide evidence using fine-grained ethnic groups.^43,44^

#### Lived experience commentary written by Lizzie Mitchell and Karen Persaud

Crucial to improving mental health service provision is tackling the persistent and wide-ranging inequalities faced by minoritized groups when accessing psychological support. With NICE guidelines recommending CBTp as a first line of intervention for psychosis and mandating access to equitable care, this study found the probability of receiving CBTp was lower among patients who were non-white, male, not in work or education, and below 26 and above 45-years-old. These findings are saddening, but also not surprising. Those of us who have used services are aware of the challenges and hurdles faced when advocating for yourself or a loved one, which can be silenced when being instantly judged on characteristics such as socioeconomic background, culture, race, age, or gender. These factors should be accepted, understood, and integrated to form a holistic treatment plan, but instead can result in being tarnished with judgement, assumptions, and unconscious biases.

The impacts of this are widespread: Patients simply being offered medication creates dependency on the system, which perpetuates the “revolving door” of being in and out of institutions. Absence of access to the right psychological intervention means patients and their carers do not benefit from an improved understanding of the illness and how to manage it, resulting in poorer mental and physical health outcomes and lower quality of life. For people declining treatment, the long-standing historical factors of consistent bias, coercion, distrust, miscommunication, misunderstanding, and lack of cultural awareness can create a barrier to willingly access services and meaningful engagement.

To close this inequalities gap, several pieces of recent research have found the need for culturally aware and responsive services, yet there seems to be a reluctancy to put this into practise. Qualitative or longitudinal studies exploring people’s experiences of being declined psychological support could be useful in exploring the reasons behind, and impact, of these statistics further. Further investigation into community and cultural resources being used would help to build resources for trusts and services seeking to redress inequalities.

We need to clearly identify the underlying reasons for inequalities in order to find solutions to remove this imbalance and provide people with the care they deserve.

*“All human beings are born free and equal in dignity and rights”* is the first declaration in the United Nations Declaration of Human Rights, and a long overdue cultural change within the mental health system is needed to reflect this.

## Data Availability

The data employed in this study are from the NCAP reports and cannot be shared by authors due privacy or ethical restrictions.

