## Supplementary Material for "Ethnic differences in receipt of psychological interventions in Early Intervention in Psychosis services in England: a cross-sectional study"

### Supplemental Methods

#### Participants: further details

We used cross-sectional data from up to three years of the National Clinical Audit of Psychosis (NCAP) commissioned across England by the Healthcare Quality Improvement Partnership (HQIP), a body working to promote quality across healthcare services.[22] Data were collected retrospectively via a case-note audit and service-level questionnaire completed by EIP teams. All NHS-funded EIP teams in England were asked to return their audit on a sample from their caseload of up to 100 people with first episode psychosis who met eligibility criteria established by the NCAP methodology.[23] If a team identified more than 100 eligible service users a random sample was selected by the NCAP. Data were collected from 162 EIP teams in all 57 eligible service providers including mental health trusts and other NHS service providers in 2018/2019, from 155 EIP teams in all 57 eligible service providers in 2019/2020, and 154 EIP teams in 55 eligible service providers in 2020/2021. Each year, data were submitted for between 97% and 99% of the total expected number of EIP service users.[24-26]

#### Outcome: further details

Data related to the receipt of CBTp and family intervention were our two main outcome variables. Receipt reflected both offer and uptake of appropriate and relevant care. We created a binary variable to indicate receipt of at least one session of each outcome in each year; in the 2018/2019 audit, CBTp and family intervention were categorized as binary variables. For the 2019/2020 and 2020/2021 audits, we categorized participants who refused, had not been offered CBTp or family intervention, or were still waiting for CBTp or family intervention as not having received CBTp or family intervention, respectively. Those who received at least one session of CBTp or family intervention were categorized as having received CBTp or family intervention, respectively.

In the sensitivity analyses (*see* “Statistical Analyses” section of the paper) we used 2019/20 and 2020/21 data to re-analyze the data to estimate any ethnic variation in “offer” (rather than receipt) of psychological interventions, categorized as offer (received, refused, waiting) versus no offer.

#### Statistical analyses: further details

We first fitted null multilevel models for each outcome including random effects. Where we found evidence of statistically significant random effects at the team level, this was retained in univariable and multivariable models. We reported intraclass correlation coefficients (ICCs) which estimate the proportion of variance in the outcome attributable to the Team level conditional on a random slope of 0 for each of the models. We reported unadjusted effect sizes for each outcome from univariable models. Our multivariable approach added age and gender to the model, followed by remaining covariates based on their lowest AIC score from univariable analyses. We reported whether fixed effects improved the model using likelihood ratio testing. The final adjusted model for the outcome receipt of "either intervention" was used for receipt of CBTp and family intervention separately to aid comparability.

### Supplemental Results

#### Descriptive statistics: further details

While 52·63% of women and 53·33% of people classified as “other gender” had received CBTp, only 44·10% of men had ($x^2(2)$ = 204·60, p<0·001). More people who were in work, education, or training had received CBTp (54·20%) compared with the proportion of those who were not (42·85%; $x^2(1)$ = 366·24, p<0·001). A greater percentage of White British people (50·07%) received CBTp compared with “any other” ethnic group (Table 1), although the proportion was similar for people of mixed ethnic groups, i.e., mixed Asian-White (49·23%), mixed Black Caribbean-White (48·92%), and mixed Black African-White (48·13%). People from Bangladeshi (38·92%), Pakistani (40·55%), and unknown (41·17%) ethnicities and were least likely to receive CBTp ($x^2(16)$ = 148.58, p<0.001).

The overall proportion of people who received family intervention in the sample was 21·18%. People receiving family intervention differed by age, occupational status, and ethnicity (p<0·001). A higher proportion of younger people received family intervention compared with those who did not receive family intervention (i.e., 26·73% of those under 26 years old received family intervention, versus 13. ·23% of those aged 56-66; $x^2(4)$ = 331·62, p<0·001). A greater percentage of those in work, education, or training received family intervention (24·73%) compared with those who were not in work, education, in training (18·82%) ($x^2(1)$ = 148·43, p<0·001). 22·49% of White British people received family intervention. A greater portion of mixed Asian-White (29·74%), mixed Black African-White (25·67%), and Chinese (25·21%) people received family intervention compared with other ethnic groups; receipt of family intervention was lowest for people of unknown ethnicities (15·11%), followed by those from “any other” ethnic background (16·98%), and Pakistani (17·42%) backgrounds ($x^2(16) = 92.85$, p<0·001.

### Sensitivity analyses on offer of interventions: further details

#### *Offer of CBTp*

Twelve out of 16 minoritized ethnic groups had lower adjusted odds of being offered CBTp than White British people ranging from 0·56 (0·43-0·75) for people of “any other Black” ethnicity to 0·75 (0·58-0·98) for people from an Indian background (Table S6). Compared to receipt (Table 2), differences in offer were marginally smaller among most ethnic groups, and failed to reach significance for Black Caribbean, mixed Black Caribbean-White, and Chinese participants (Table S6). Contrarily, differences were marginally increased for those of White Irish ethnicity and reached significance for people from a mixed Asian-White background (aOR: 0·51; 95%CI: 0·51-0·82).

#### *Offer of family intervention*

Five out of 16 minoritized ethnic groups had lower adjusted odds of being offered family intervention ranging from 0·65 (95%CI: 0·56-0·74) for Black African to 0·79 (95%CI: 0·65-0·97) for Black Caribbean participants (Table S6). Compared to receipt (Table 2), differences in offer were marginally smaller for people of Black African, Black Caribbean, and “any other” ethnicity and failed to reach significance for those from “any other Black” background (Table S6). Contrary to receipt, Indian people had higher adjusted odds of being offered family intervention compared to White British people (95%CI: 1·26; 1·01-1·56).

#### *Offer of either intervention*

Eight out of 16 minoritized ethnic groups had lower adjusted odds of being offered either intervention with odds ranging from 0·46 (95%CI: 0·24-0·87) for Chinese participants to 0·74 (95%CI: 0·60-0·92) for people of unknown ethnicity. Six of the ethnic groups with lower adjusted odds of receiving either intervention showed no differences in being offered either of the two interventions (Table 2 and S6). Contrarily, decreased adjusted odds for those from mixed Asian-White background reached significance 0·50 (95%CI: 0·29-0·87).

### Supplemental Figures and Tables

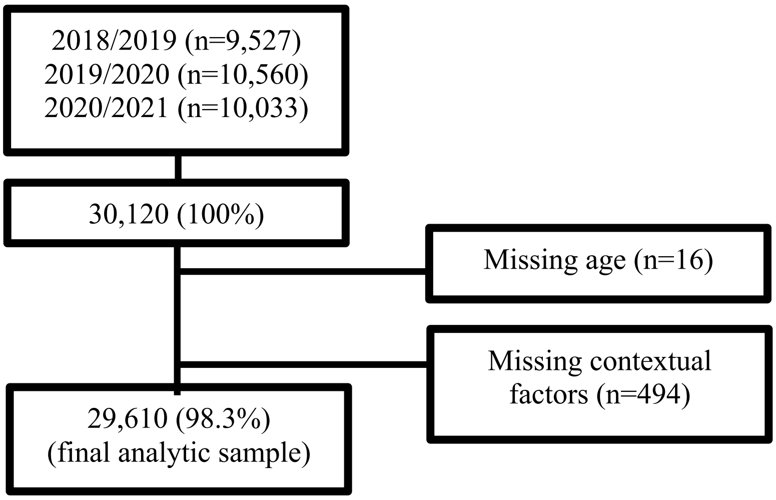


**Figure 1: PRISMA flow chart of values dropped from the final analytic sample.**

**Table S1: Sociodemographic characteristics, ethnicities, and outcomes, by included and excluded participants**

| | Total | Missing | Not missing | $x^2$ (df) | $p(x^2)$ |
|---|---|---|---|---|---|
| **Ethnicity** | | | | 353·54 (5)* | <0·001 |
| White British | 17,266 | 111 (0·64) | 17,155 (99·36) | | |
| White Irish | 167 | 6 (3·59) | 161 (96·41) | | |
| Any other White background | 1,926 | 63 (3·27) | 1,863 (96·73) | | |
| Black African | 2,145 | 71 (3·31) | 2,074 (96·69) | | |
| Black Caribbean | 979 | 62 (6·33) | 917 (93·67) | | |
| Mixed Black African-White | 190 | 3 (1·58) | 187 (98·42) | | |
| Mixed Black Caribbean-White | 377 | 5 (1·33) | 372 (98·67) | | |
| Any other Black background | 676 | 37 (5·47) | 639 (94·53) | | |
| Bangladeshi | 596 | 5 (0·84) | 591 (99·16) | | |
| Indian | 737 | 28 (3·80) | 709 (96·20) | | |
| Pakistani | 1,371 | 22 (1·60) | 1,349 (98·40) | | |
| Chinese | 120 | 1 (0·83) | 119 (99·17) | | |
| Mixed Asian-White | 201 | 6 (2·99) | 195 (97·01) | | |
| Any other Asian background | 1,002 | 33 (3·29) | 969 (96·71) | | |
| Any other mixed background | 465 | 15 (3·23) | 450 (96·77) | | |
| Any other ethnic background | 832 | 31 (3·73) | 801 (96·27) | | |
| Unknown or refused | 1,070 | 11 (1·03) | 1,059 (98·97) | | |
| **Gender** | | | | 1·42 (na) | 0·616** |
| Male | 18,546 | 304 (1·64) | 18,242 (98·36) | | |
| Female | 11,544 (100) | 206 (1·78) | 11,338 (98·22) | | |
| Other | 30 (100) | 0 (0·00) | 30 (100·00) | | |
| **Age category** | | | | 8·97 (4) | 0·062 |
| <26 | 10,325 | 187 (1·81) | 10,138 (98·19) | | |
| 26-35 | 10,664 | 184 (1·73) | 10,480 (98·27) | | |
| 36-45 | 4,805 | 73 (1·52) | 4,732 (98·48) | | |
| 46-55 | 2,789 | 33 (1·18) | 2,756 (98·82) | | |
| 56-66 | 1,521 | 17 (1·12) | 1,504 (98·88) | | |
| **Occupational status** | | | | 0·37 (1) | 0·542 |
| In work, education, or training | 12,008 | 210 (1·75) | 11,798 (98·25) | | |
| Not in work, education, or training | 18,112 | 300 (1·66) | 17,812 (98·34) | | |
| **CBTp** | | | | 0.32 (1) | 0·574 |
| Not received | 15,845 | 262 (1·65) | 15,583 (98·35) | | |
| Received | 14,275 | 248 (1·74) | 14,027 (98·26) | | |
| **Family intervention** | | | | 0.18 (1) | 0·668 |
| Not received | 23,737 | 398 (1·68) | 23,339 (98·32) | | |
| Received | 6,383 | 112 (1·75) | 6,271 (98·25) | | |
| **Either intervention** | | | | 0.01 (1) | 0·911 |
| Not received | 13,510 | 230 (1·70) | 13,280 (98·30) | | |
| Received | 16,610 | 280 (1·69) | 16,330 (98·31) | | |

*for White British vs White other, Black, Asian, mixed and other, and unknown ethnicities. Fisher's exact would not converge on 17 categories.
**based on Fisher's exact.

**Table S2: Sociodemographic characteristics and ethnicities, by offer of CBTp, family intervention, and either intervention for years 2019-2020 and 2020-2021**

| Characteristics | Full sample | CBTp | | | | Family intervention | | | | Either intervention | | | |
|---|---|---|---|---|---|---|---|---|---|---|---|---|---|
| | | Not offered | Offered | | | Not offered | Offered | | | Not offered | Offered | | |
| | *N* (100%) | *N* (%) | *N* (%) | $x^2$ (df) | p($x^2$) | *N* (%) | *N* (%) | $x^2$ (df) | p($x^2$) | *N* (%) | *N* (%) | $x^2$ (df) | p($x^2$) |
| **Total** | 20,366 | 3,897 (19·13) | 16,469 (80·87) | | | 7,685 (37·73) | 12,681 (62·27) | | | 2,525 (12·40) | 17,841 (87·60) | | |
| **Gender** | | | | 43·82 (2) | <0·001 | | | 3·51 (2) | 0·173 | | | 27·97 (2) | <0·001 |
| Male | 12,527 | 2,569 (20·51) | 9,958 (79·49) | | | 4,790 (38·24) | 7,737 (61·76) | | | 1,674 (13·36) | 10,853 (86·64) | | |
| Female | 7,815 | 1,320 (16·89) | 6,495 (83·11) | | | 2,886 (36·93) | 4,929 (63·07) | | | 848 (10·85) | 6,967 (89·15) | | |
| Other | 24 | 8 (33·33) | 16 (66·67) | | | 9 (37·50) | 15 (62·50) | | | 3 (12·50) | 21 (87·50) | | |
| **Age** | | | | 30·76 (4) | <0·001 | | | 90·50 (4) | <0·001 | | | 6·65 (4) | 0·155 |
| <26 | 6,570 | 1,384 (21·07) | 5,186 (78·93) | | | 2,192 (33·36) | 4,378 (66·64) | | | 844 (12·85) | 5,726 (87·15) | | |
| 26-35 | 7,097 | 1,355 (19·09) | 5,742 (80·91) | | | 2,730 (38·47) | 4,367 (61·53) | | | 903 (12·72) | 6,194 (87·28) | | |
| 36-45 | 3,458 | 592 (17·12) | 2,866 (82·88) | | | 1,415 (40·92) | 2,043 (59·08) | | | 390 (11·28) | 3,068 (88·72) | | |
| 46-55 | 2,056 | 359 (17·46) | 1,697 (82·54) | | | 855 (41·59) | 1,201 (58·41) | | | 242 (11·77) | 1,814 (88·23) | | |
| 56-66 | 1,185 | 207 (17·47) | 978 (82·53) | | | 493 (41·60) | 692 (58·40) | | | 146 (12·32) | 1,039 (87·68) | | |
| **Occupational status** | | | | 35·34 (1) | <0·001 | | | 67·89 (1) | <0·001 | | | 68·20 (1) | <0·001 |
| In work, education, or training | 8,182 | 1,402 (17·14) | 6,780 (82·86) | | | 2,808 (34·32) | 5,374 (65·68) | | | 824 (10·07) | 7,358 (89·93) | | |
| Not in work, education, or training | 12,184 | 2,495 (20·48) | 9,689 (79·52) | | | 4,877 (40·03) | 7,307 (59·97) | | | 1,701 (13·96) | 10,483 (86·04) | | |
| **Ethnicity** | | | | 83·17 (16) | <0·001 | | | 115·87 (16) | <0·001 | | | 106·87 (16) | <0·001 |
| White British | 11,699 | 2,139 (18·28) | 9,560 (81·72) | | | 4,235 (36·20) | 7,464 (63·80) | | | 1,328 (11·35) | 10,371 (88·65) | | |
| White Irish | 110 | 30 (27·27) | 80 (72·73) | | | 44 (40·00) | 66 (60·00) | | | 19 (17·27) | 91 (82·73) | | |
| Any other White background | 1,290 | 229 (17·75) | 1,061 (82·25) | | | 488 (37·83) | 802 (62·17) | | | 158 (12·25) | 1,132 (87·75) | | |
| Black African | 1,439 | 249 (17·30) | 1,190 (82·70) | | | 595 (41·35) | 844 (58·65) | | | 183 (12·72) | 1,256 (87·28) | | |
| Black Caribbean | 632 | 116 (18·35) | 516 (81·65) | | | 249 (39·40) | 383 (60·60) | | | 70 (11·08) | 562 (88·92) | | |
| Mixed Black African-White | 122 | 26 (21·31) | 96 (78·69) | | | 42 (34·43) | 80 (65·57) | | | 16 (13·11) | 106 (86·89) | | |
| Mixed Black Caribbean-White | 235 | 40 (17·02) | 195 (82·98) | | | 78 (33·19) | 157 (66·81) | | | 27 (11·49) | 208 (88·51) | | |
| Any other Black background | 399 | 87 (21·80) | 312 (78·20) | | | 159 (39·85) | 240 (60·15) | | | 59 (14·79) | 340 (85·21) | | |
| Bangladeshi | 398 | 74 (18·59) | 324 (81·41) | | | 143 (35·93) | 255 (64·07) | | | 52 (13·07) | 346 (86·93) | | |
| Indian | 525 | 102 (19·43) | 423 (80·57) | | | 171 (32·57) | 354 (67·43) | | | 52 (9·90) | 473 (90·10) | | |
| Pakistani | 913 | 198 (21·69) | 715 (78·31) | | | 362 (39·65) | 551 (60·35) | | | 126 (13·80) | 787 (86·20) | | |
| Chinese | 86 | 19 (22·09) | 67 (77·91) | | | 28 (32·56) | 58 (67·44) | | | 16 (18·60) | 70 (81·40) | | |
| Mixed Asian-White | 128 | 30 (23·44) | 98 (76·56) | | | 42 (32·81) | 86 (67·19) | | | 20 (15·63) | 108 (84·38) | | |
| Any other Asian background | 661 | 126 (19·06) | 535 (80·94) | | | 241 (36·46) | 420 (63·54) | | | 80 (12·10) | 581 (87·90) | | |
| Any other mixed background | 332 | 69 (20·78) | 263 (79·22) | | | 139 (41·87) | 193 (58·13) | | | 47 (14·16) | 285 (85·84) | | |

| | | | | | | | |
|---|---|---|---|---|---|---|---|
| Any other ethnic background | 566 | 118 (20.85) | 448 (79.15) | 234 (41·34) | 332 (58·66) | 87 (15·37) | 479 (84·63) |
| Unknown or refused | 831 | 245 (29.48) | 586 (70.52) | 435 (52·35) | 396 (47·65) | 185 (22·26) | 646 (77·74) |

**Table S3: Association between ethnicities and receipt of CBTp, family intervention, and either intervention in 2018-2019**

| | CBTp | | Family intervention | | Either intervention* | |
|---|---|---|---|---|---|---|
| | OR (95%CI:) | aOR (95%CI:) | OR (95%CI:) | aOR (95%CI:) | OR (95%CI:) | aOR (95%CI:) |
| **Fixed Effects** | | | | | | |
| *Ethnicity* | | | | | | |
| White British | 1 | 1 | 1 | 1 | 1 | 1 |
| White Irish | 0·69 (0·38-1·26) | 0·74 (0·41-1·35) | 1·01 (0·49-2·06) | 1·05 (0·51-2·15) | 0·67 (0·37-1·23) | 0·71 (0·39-1·30) |
| Any other White background | **0·65 (0·54-0·79)** | **0·61 (0·50-0·74)** | 0·85 (0·67-1·08) | 0·83 (0·65-1·05) | **0·71 (0·58-0·86)** | **0·67 (0·55-0·81)** |
| Black African | **0·61 (0·50-0·74)** | **0·59 (0·48-0·71)** | **0·67 (0·52-0·85)** | **0·62 (0·49-0·80)** | **0·61 (0·50-0·73)** | **0·58 (0·47-0·70)** |
| Black Caribbean | **0·57 (0·43-0·75)** | **0·56 (0·43-0·74)** | 0·82 (0·59-1·14) | 0·80 (0·57-1·11) | **0·62 (0·47-0·81)** | **0·60 (0·46-0·79)** |
| Mixed Black African-White | 0·76 (0·45-1·29) | 0·76 (0·44-1·28) | 0·99 (0·54-1·83) | 0·85 (0·45-1·58) | 0·68 (0·40-1·15) | 0·65 (0·38-1·10) |
| Mixed Black Caribbean-White | 1·07 (0·74-1·56) | 1·07 (0·74-1·56) | 1·10 (0·71-1·69) | 1·02 (0·66-1·58) | 1·15 (0·78-1·68) | 1·11 (0·76-1·64) |
| Any other Black background | **0·62 (0·46-0·83)** | **0·61 (0·45-0·82)** | 0·74 (0·52-1·05) | **0·67 (0·47-0·97)** | **0·69 (0·51-0·93)** | **0·66 (0·49-0·90)** |
| Bangladeshi | **0·44 (0·31-0·63)** | **0·42 (0·30-0·60)** | 1·29 (0·87-1·91) | 1·19 (0·80-1·77) | **0·61 (0·44-0·86)** | **0·58 (0·41-0·81)** |
| Indian | **0·65 (0·47-0·90)** | **0·61 (0·44-0·85)** | 1·31 (0·90-1·92) | 1·27 (0·87-1·87) | 0·78 (0·57-1·08) | 0·74 (0·53-1·03) |
| Pakistani | **0·67 (0·53-0·84)** | **0·64 (0·51-0·81)** | 0·96 (0·72-1·28) | 0·91 (0·68-1·22) | **0·74 (0·59-0·92)** | **0·71 (0·56-0·89)** |
| Chinese | 0·55 (0·26-1·16) | **0·46 (0·22-0·97)** | 0·96 (0·41-2·25) | 0·89 (0·37-2·11) | 0·62 (0·30-1·28) | 0·52 (0·25-1·10) |
| Mixed Asian-White | 0·98 (0·59-1·64) | 0·96 (0·57-1·61) | 1·55 (0·89-2·72) | 1·38 (0·78-2·45) | 1·22 (0·72-2·08) | 1·16 (0·68-1·99) |
| Any other Asian background | **0·56 (0·43-0·73)** | **0·55 (0·43-0·72)** | 1·10 (0·81-1·50) | 1·09 (0·80-1·49) | **0·69 (0·54-0·90)** | **0·69 (0·53-0·89)** |
| Any other mixed background | **0·65 (0·43-0·97)** | **0·62 (0·41-0·93)** | 0·84 (0·51-1·38) | 0·78 (0·47-1·29) | **0·65 (0·44-0·97)** | **0·62 (0·41-0·93)** |
| Any other ethnic background | **0·66 (0·50-0·88)** | **0·67 (0·50-0·90)** | **0·66 (0·45-0·98)** | 0·68 (0·45-1·00) | **0·62 (0·46-0·82)** | **0·63 (0·47-0·84)** |
| Unknown or refused | **0·70 (0·51-0·95)** | **0·69 (0·50-0·95)** | 0·78 (0·50-1·23) | 0·78 (0·49-1·23) | **0·68 (0·50-0·92)** | **0·67 (0·49-0·92)** |
| *Gender* | | | | | | |
| Female | **1·42 (1·30-1·56)** | **1·42 (1·29-1·56)** | 1·02 (0·91-1·14) | 1·10 (0·98-1·23) | **1·37 (1·25-1·50)** | **1·4 (1·27-1·53)** |
| Other/undefined | **9·36 (1·04-83·90)** | **9·18 (1·02-82·20)** | 0·86 (0·09-8·36) | 0·77 (0·08-7·30) | omitted | omitted |
| Male | 1 | 1 | 1 | 1 | 1 | 1 |
| *Age* | | | | | | |
| <26 | 1·02 (0·92-1·13) | 0·96 (0·87-1·07) | **1·66 (1·47-1·88)** | **1·60 (1·41-1·82)** | 1·15 (1·04-1·27) | 1·09 (0·99-1·21) |
| 26-35 | 1 | 1 | 1 | 1 | 1 | 1 |
| 36-45 | 1·12 (0·98-1·29) | 1·09 (0·95-1·26) | **0·80 (0·66-0·96)** | **0·78 (0·65-0·94)** | 1·00 (0·87-1·15) | 0·97 (0·84-1·11) |
| 46-55 | 0·93 (0·78-1·11) | 0·87 (0·73-1·04) | **0·75 (0·59-0·96)** | **0·74 (0·58-0·95)** | **0·83 (0·69-0·99)** | **0·77 (0·65-0·93)** |
| 56-66 | 0·82 (0·64-1·05) | **0·72 (0·56-0·93)** | **0·43 (0·29-0·64)** | **0·41 (0·28-0·61)** | **0·71 (0·55-0·91)** | **0·63 (0·49-0·81)** |
| *Occupational status* | | | | | | |
| In work, education, or training | **1·68 (1·53-1·84)** | **1·67 (1·52-1·83)** | **1·46 (1·31-1·63)** | **1·35 (1·20-1·51)** | **1·62 (1·48-1·78)** | **1·57 (1·42-1·72)** |
| Not in work, education, or training | 1 | 1 | 1 | 1 | 1 | 1 |
| *Caseload size of care coordinators* | 1·00 (0·98-1·03) | 1·01 (0·99-1·03) | 1·00 (0·97-1·02) | 1·00 (0·97-1·03) | 1·00 (0·98-1·02) | 1·00 (0·98-1·03) |
| *Inequalities strategies* | | | | | | |
| Yes | 0·98 (0·74-1·31) | 1·00 (0·74-1·35) | 1·17 (0·82-1·68) | 1·12 (0·77-1·62) | 1·02 (0·78-1·35) | 1·02 (0·77-1·36) |
| No | 1 | 1 | 1 | 1 | 1 | 1 |
| **Random Part of the Model** | | | | | | |
| *ICC* | | | | | | |
| Null | **0·18 (0·14-0·22)** | **0·18 (0·14-0·22)** | **0·25 (0·20-0·31)** | **0·25 (0·20-0·31)** | **0·16 (0·13-0·20)** | **0·16 (0·13-0·20)** |
| Unadjusted/adjusted | **0·19 (0·15-0·24)** | **0·19 (0·15-0·24)** | **0·25 (0·20-0·31)** | **0·26 (0·21-0·32)** | **0·17 (0·13-0·21)** | 1·02 (0·77-1·36) |

OR: Odds ratio; 95%CI: 95% confidence interval. P-values ($p<0·05$) in bold.

*n=9,238 (6 observations of "other gender" omitted)

[1]Univariable regression analyses including the outcome and random effects.
[2]Full adjusted model including fixed effects for ethnicity, gender, age, occupational status, caseload size of care coordinators, and inequalities strategies, and random effects for EIP teams and year of data collection.
[3]Caseload size is scaled as the odds associated with one extra patient per care coordinator (i.e., increased caseload size of care coordinators is associated with lower odds of receipt of either psychological intervention).
[4]ICC reported from an unadjusted model including ethnicity as the single covariate.

**Table S4: Association between ethnicities and receipt of CBTp, family intervention, and either intervention in 2019-2020**

| | CBTp | | Family intervention | | Either intervention | |
|---|---|---|---|---|---|---|
| | OR (95%CI:) | aOR (95%CI:) | OR (95%CI:) | aOR (95%CI:) | OR (95%CI:) | aOR (95%CI:) |
| **Fixed Effects** | | | | | | |
| *Ethnicity* | | | | | | |
| White British | 1 | 1 | 1 | 1 | 1 | 1 |
| White Irish | 0·73 (0·41-1·31) | 0·75 (0·42-1·35) | 1·07 (0·54-2·13) | 1·13 (0·57-2·25) | 0·67 (0·37-1·20) | 0·68 (0·38-1·23) |
| Any other White background | **0·69 (0·58-0·82)** | **0·65 (0·54-0·77)** | **0·60 (0·47-0·76)** | **0·59 (0·46-0·75)** | **0·65 (0·54-0·77)** | **0·61 (0·51-0·73)** |
| Black African | **0·53 (0·45-0·63)** | **0·51 (0·43-0·61)** | **0·66 (0·53-0·83)** | **0·63 (0·50-0·78)** | **0·52 (0·44-0·62)** | **0·49 (0·41-0·59)** |
| Black Caribbean | **0·60 (0·47-0·76)** | **0·60 (0·47-0·77)** | **0·66 (0·49-0·90)** | **0·64 (0·47-0·88)** | **0·55 (0·43-0·69)** | **0·54 (0·43-0·70)** |
| Mixed Black African-White | 0·71 (0·43-1·19) | 0·68 (0·41-1·15) | 1·03 (0·57-1·85) | 0·90 (0·50-1·63) | 0·81 (0·48-1·37) | 0·76 (0·45-1·29) |
| Mixed Black Caribbean-White | **0·67 (0·46-0·97)** | **0·68 (0·47-1·00)** | **0·59 (0·35-0·99)** | **0·54 (0·32-0·91)** | **0·60 (0·41-0·87)** | **0·59 (0·41-0·87)** |
| Any other Black background | **0·61 (0·46-0·82)** | **0·61 (0·45-0·83)** | 0·69 (0·47-1·02) | **0·64 (0·43-0·95)** | **0·59 (0·44-0·79)** | **0·58 (0·43-0·79)** |
| Bangladeshi | **0·38 (0·27-0·52)** | **0·35 (0·25-0·49)** | 1·02 (0·69-1·51) | 0·94 (0·64-1·40) | **0·43 (0·31-0·60)** | **0·39 (0·28-0·55)** |
| Indian | **0·70 (0·53-0·92)** | **0·67 (0·50-0·88)** | 1·33 (0·97-1·84) | 1·31 (0·94-1·81) | 0·88 (0·67-1·16) | 0·85 (0·64-1·13) |
| Pakistani | **0·49 (0·40-0·61)** | **0·47 (0·38-0·59)** | 0·82 (0·62-1·08) | 0·78 (0·59-1·03) | **0·49 (0·40-0·61)** | **0·47 (0·38-0·58)** |
| Chinese | 0·81 (0·40-1·63) | 0·70 (0·34-1·42) | 1·00 (0·43-2·33) | 0·95 (0·40-2·25) | 0·67 (0·33-1·36) | 0·58 (0·28-1·19) |
| Mixed Asian-White | 0·75 (0·44-1·28) | 0·70 (0·40-1·20) | 1·33 (0·73-2·44) | 1·12 (0·61-2·08) | 0·73 (0·43-1·24) | 0·65 (0·37-1·11) |
| Any other Asian background | **0·60 (0·47-0·76)** | **0·58 (0·45-0·73)** | 0·94 (0·70-1·26) | 0·90 (0·67-1·21) | **0·59 (0·47-0·75)** | **0·56 (0·44-0·72)** |
| Any other mixed background | 0·87 (0·62-1·21) | 0·85 (0·60-1·18) | 0·69 (0·45-1·07) | **0·62 (0·40-0·96)** | 0·72 (0·51-1·01) | **0·68 (0·48-0·95)** |
| Any other ethnic background | **0·60 (0·46-0·77)** | **0·60 (0·46-0·78)** | 0·76 (0·54-1·06) | 0·75 (0·54-1·05) | **0·56 (0·43-0·72)** | **0·55 (0·43-0·72)** |
| Unknown or refused | **0·71 (0·56-0·90)** | **0·70 (0·55-0·89)** | 0·77 (0·55-1·06) | 0·73 (0·52-1·01) | **0·67 (0·53-0·85)** | **0·65 (0·51-0·82)** |
| *Gender* | | | | | | |
| Female | **1·47 (1·35-1·59)** | **1·49 (1·37-1·62)** | 1·04 (0·94-1·15) | **1·13 (1·02-1·25)** | **1·41 (1·30-1·54)** | **1·48 (1·36-1·62)** |
| Other | 1·67 (0·45-6·19) | 1·63 (0·44-6·05) | 1·23 (0·23-6·60) | 1·12 (0·21-6·15) | 3·23 (0·77-13·53) | 3·08 (0·73-12·94) |
| Male | 1 | 1 | 1 | 1 | 1 | 1 |
| *Age* | | | | | | |
| <26 | 0·93 (0·84-1·02) | **0·86 (0·78-0·95)** | 1·42 (1·26-1·60) | **1·36 (1·21-1·54)** | 1·04 (0·95-1·15) | 0·98 (0·88-1·08) |
| 26-35 | 1 | **1** | 1 | 1 | 1 | 1 |
| 36-45 | **0·86 (0·76-0·97)** | **0·82 (0·73-0·93)** | **0·76 (0·65-0·89)** | **0·74 (0·64-0·87)** | **0·81 (0·72-0·91)** | **0·77 (0·68-0·87)** |
| 46-55 | 0·95 (0·82-1·10) | **0·83 (0·72-0·97)** | **0·71 (0·58-0·86)** | **0·66 (0·55-0·81)** | **0·84 (0·72-0·97)** | **0·74 (0·63-0·86)** |
| 56-66 | 0·88 (0·73-1·07) | **0·74 (0·61-0·90)** | **0·46 (0·34-0·60)** | **0·42 (0·32-0·56)** | **0·75 (0·62-0·91)** | **0·63 (0·52-0·76)** |
| *Occupational status* | | | | | | |
| In work, education, or training | **1·62 (1·49-1·76)** | **1·62 (1·48-1·76)** | **1·37 (1·24-1·52)** | **1·27 (1·14-1·41)** | **1·62 (1·48-1·76)** | **1·57 (1·44-1·71)** |
| Not work, education, or training | 1 | 1 | 1 | 1 | 1 | 1 |
| *Caseload size of care coordinators* | 0·99 (0·97-1·01) | 1·00 (0·97-1·02) | 0·98 (0·95-1·01) | 0·98 (0·95-1·01) | 0·98 (0·96-1·01) | 0·99 (0·97-1·02) |
| *Inequalities strategies* | | | | | | |
| Yes | 0·92 (0·73-1·16) | 0·94 (0·73-1·21) | 0·87 (0·65-1·17) | 0·86 (0·63-1·17) | 0·90 (0·71-1·13) | 0·92 (0·71-1·18) |
| No | 1 | 1 | 1 | 1 | 1 | 1 |
| **Random Part of the Model** | | | | | | |
| *ICC* | | | | | | |
| Null | **0·12 (0·09-0·15)** | **0·12 (0·09-0·15)** | **0·18 (0·14-0·22)** | **0·18 (0·14-0·22)** | **0·12 (0·09-0·15)** | **0·12 (0·09-0·15)** |
| Unadjusted/adjusted | **0·13 (0·10-0·17)** | **0·14 (0·11-0·17)** | **0·18 (0·14-0·22)** | **0·19 (0·15-0·24)** | **0·13 (0·10-0·16)** | **0·14 (0·11-0·17)** |

OR: Odds ratio; 95%CI: 95% confidence interval. P-values (p<0·05) in bold.

[1]Univariable regression analyses including the outcome and random effects.

[2]Full adjusted model including fixed effects for ethnicity, gender, age, occupational status, caseload size of care coordinators, and inequalities strategies, and random effects for EIP teams and year of data collection.
[3]Caseload size is scaled as the odds associated with one extra patient per care coordinator (i.e., increased caseload size of care coordinators is associated with lower odds of receipt of either psychological intervention).
[4]ICC reported from an unadjusted model including ethnicity as the single covariate.

**Table S5: Association between ethnicities and receipt of CBTp, family intervention, and either intervention in 2020-2021**

| | CBTp | | Family intervention | | Either intervention | |
|---|---|---|---|---|---|---|
| | OR (95%CI:) | aOR (95%CI:) | OR (95%CI:) | aOR (95%CI:) | OR (95%CI:) | aOR (95%CI:) |
| **Fixed Effects** | | | | | | |
| *Ethnicity* | | | | | | |
| White British | 1 | 1 | 1 | 1 | 1 | 1 |
| White Irish | **0·56 (0·32-0·99)** | 0·59 (0·34-1·04) | 0·62 (0·31-1·26) | 0·66 (0·32-1·34) | **0·52 (0·30-0·89)** | **0·54 (0·32-0·94)** |
| Any other White background | **0·64 (0·53-0·76)** | **0·59 (0·49-0·71)** | 0·84 (0·68-1·04) | 0·82 (0·66-1·02) | **0·65 (0·54-0·77)** | **0·60 (0·50-0·72)** |
| Black African | **0·58 (0·48-0·69)** | **0·54 (0·44-0·65)** | **0·65 (0·52-0·83)** | **0·60 (0·47-0·76)** | **0·58 (0·48-0·70)** | **0·53 (0·44-0·64)** |
| Black Caribbean | **0·64 (0·49-0·84)** | **0·65 (0·50-0·85)** | **0·61 (0·43-0·86)** | **0·61 (0·43-0·87)** | **0·59 (0·46-0·77)** | **0·59 (0·45-0·78)** |
| Mixed Black African-White | 0·91 (0·52-1·60) | 0·90 (0·51-1·59) | 1·21 (0·64-2·32) | 1·06 (0·55-2·03) | 1·07 (0·61-1·90) | 1·01 (0·57-1·79) |
| Mixed Black Caribbean-White | 0·69 (0·46-1·05) | 0·71 (0·47-1·07) | 0·94 (0·58-1·54) | 0·85 (0·52-1·39) | 0·72 (0·48-1·08) | 0·70 (0·46-1·05) |
| Any other Black background | **0·48 (0·34-0·66)** | **0·45 (0·33-0·63)** | **0·61 (0·41-0·92)** | **0·59 (0·39-0·89)** | **0·42 (0·31-0·58)** | **0·40 (0·29-0·55)** |
| Bangladeshi | **0·46 (0·33-0·64)** | **0·43 (0·31-0·61)** | 0·71 (0·46-1·08) | 0·65 (0·42-1·01) | **0·53 (0·38-0·73)** | **0·49 (0·35-0·69)** |
| Indian | **0·64 (0·49-0·84)** | **0·60 (0·46-0·79)** | 1·01 (0·73-1·39) | 1·03 (0·75-1·43) | **0·72 (0·55-0·94)** | **0·69 (0·53-0·90)** |
| Pakistani | **0·59 (0·47-0·74)** | **0·57 (0·45-0·71)** | 0·91 (0·69-1·21) | 0·88 (0·66-1·17) | **0·65 (0·52-0·81)** | **0·62 (0·50-0·78)** |
| Chinese | **0·51 (0·28-0·93)** | **0·47 (0·25-0·86)** | 1·04 (0·53-2·01) | 1·22 (0·62-2·38) | 0·64 (0·35-1·17) | 0·63 (0·35-1·15) |
| Mixed Asian-White | 0·93 (0·56-1·55) | 0·90 (0·54-1·52) | 1·14 (0·64-2·04) | 0·98 (0·55-1·76) | 1·19 (0·70-2·03) | 1·11 (0·65-1·89) |
| Any other Asian background | **0·69 (0·54-0·89)** | **0·66 (0·51-0·84)** | **0·67 (0·49-0·92)** | **0·63 (0·46-0·87)** | **0·65 (0·51-0·83)** | **0·62 (0·48-0·79)** |
| Any other mixed background | 0·74 (0·53-1·03) | **0·69 (0·49-0·96)** | 0·93 (0·63-1·39) | 0·79 (0·53-1·18) | 0·85 (0·61-1·18) | 0·76 (0·55-1·06) |
| Any other ethnic background | **0·51 (0·39-0·67)** | **0·50 (0·38-0·65)** | **0·61 (0·43-0·86)** | **0·57 (0·41-0·81)** | **0·52 (0·40-0·67)** | **0·50 (0·39-0·65)** |
| Unknown or refused | **0·68 (0·54-0·86)** | **0·66 (0·52-0·83)** | 0·84 (0·62-1·13) | 0·78 (0·57-1·05) | **0·70 (0·56-0·88)** | **0·67 (0·53-0·84)** |
| *Gender* | | | | | | |
| Female | **1·36 (1·25-1·48)** | **1·37 (1·25-1·50)** | 0·95 (0·86-1·06) | 1·02 (0·91-1·14) | **1·26 (1·15-1·37)** | **1·30 (1·19-1·42)** |
| Other | 0·96 (0·32-2·93) | 0·82 (0·27-2·55) | 1·29 (0·34-4·86) | 0·90 (0·25-3·32) | 1·16 (0·38-3·59) | 0·89 (0·29-2·80) |
| Male | 1 | 1 | 1 | 1 | 1 | 1 |
| *Age* | | | | | | |
| <26 | 0·99 (0·89-1·09) | 0·93 (0·83-1·03) | **1·63 (1·44-1·84)** | **1·57 (1·39-1·78)** | 1·15 (1·04-1·28) | 1·09 (0·98-1·21) |
| 26-35 | 1 | 1 | 1 | 1 | 1 | 1 |
| 36-45 | 1·05 (0·93-1·19) | 1·03 (0·91-1·17) | 0·86 (0·73-1·00) | 0·85 (0·73-1·00) | 0·98 (0·86-1·11) | 0·96 (0·85-1·09) |
| 46-55 | 0·99 (0·85-1·15) | 0·89 (0·76-1·04) | **0·72 (0·59-0·88)** | **0·70 (0·57-0·85)** | 0·90 (0·77-1·05) | **0·82 (0·70-0·95)** |
| 56-66 | 0·87 (0·73-1·05) | **0·77 (0·64-0·93)** | **0·68 (0·53-0·86)** | **0·65 (0·50-0·83)** | **0·78 (0·65-0·93)** | **0·69 (0·57-0·83)** |
| *Occupational status* | | | | | | |
| In work, education, or training | **1·63 (1·49-1·78)** | **1·64 (1·50-1·79)** | **1·51 (1·36-1·67)** | **1·41 (1·26-1·56)** | **1·65 (1·51-1·80)** | **1·61 (1·48-1·76)** |
| Not in work, education, or training | 1 | 1 | 1 | 1 | 1 | 1 |
| *Caseload size of care coordinators* | 0·98 (0·96-1·00) | 0·99 (0·97-1·01) | **0·97 (0·94-0·99)** | **0·97 (0·95-1·00)** | **0·97 (0·95-0·99)** | **0·98 (0·96-1·00)** |
| *Inequalities* | | | | | | |
| Yes | 1·19 (0·94-1·52) | 1·23 (0·95-1·60) | 1·18 (0·88-1·58) | 1·1 (0·82-1·49) | 1·22 (0·97-1·54) | 1·23 (0·96-1·56) |
| No | 1 | 1 | 1 | 1 | 1 | 1 |
| **Random Part of the Model** | | | | | | |
| *ICC* | | | | | | |
| Null | **0·12 (0·09-0·16)** | **0·12 (0·09-0·16)** | **0·16 (0·13-0·21)** | **0·16 (0·13-0·21)** | **0·11 (0·08-0·14)** | **0·11 (0·08-0·14)** |
| Unadjusted/adjusted | **0·13 (0·10-0·17)** | **0·13 (0·10-0·17)** | **0·17 (0·13-0·21)** | **0·17 (0·13-0·21)** | **0·12 (0·09-0·15)** | **0·11 (0·09-0·15)** |

OR: Odds ratio; 95%CI: 95% confidence interval. P-values (p<0·05) in bold.

[1]Univariable regression analyses including the outcome and random effects.

[2]Full adjusted model including fixed effects for ethnicity, gender, age, occupational status, caseload size of care coordinators, and inequalities strategies, and random effects for EIP teams and year of data collection.
[3]Caseload size is scaled as the odds associated with one extra patient per care coordinator (i.e., increased caseload size of care coordinators is associated with lower odds of receipt of either psychological intervention).
[4]ICC reported from an unadjusted model including ethnicity as the single covariate.

**Table S6: Association between ethnicities and offer of CBTp, family intervention, and either intervention**

| | CBTp | | Family intervention | | Either intervention | |
|---|---|---|---|---|---|---|
| | OR (95%CI:) | aOR (95%CI:) | OR (95%CI:) | aOR (95%CI:) | OR (95%CI:) | aOR (95%CI:) |
| **Fixed Effects** | | | | | | |
| *Ethnicity* | | | | | | |
| White British | 1 | 1 | 1 | 1 | 1 | 1 |
| White Irish | **0·51 (0·32-0·82)** | **0·51 (0·32-0·83)** | 0·95 (0·61-1·48) | 0·97 (0·62-1·50) | 0·59 (0·34-1·02) | 0·60 (0·34-1·03) |
| Any other White background | **0·68 (0·57-0·81)** | **0·65 (0·54-0·78)** | **0·72 (0·63-0·83)** | **0·70 (0·61-0·81)** | **0·62 (0·51-0·76)** | **0·59 (0·48-0·72)** |
| Black African | **0·67 (0·56-0·80)** | **0·66 (0·55-0·78)** | **0·69 (0·60-0·79)** | **0·65 (0·56-0·74)** | **0·63 (0·52-0·77)** | **0·61 (0·50-0·75)** |
| Black Caribbean | **0·78 (0·61-0·99)** | 0·78 (0·62-1·02) | **0·81 (0·66-0·99)** | **0·79 (0·65-0·97)** | 0·90 (0·67-1·21) | 0·93 (0·69-1·24) |
| Mixed Black African-White | 0·72 (0·43-1·20) | 0·75 (0·45-1·26) | 0·99 (0·64-1·52) | 0·90 (0·58-1·38) | 0·86 (0·48-1·54) | 0·86 (0·48-1·56) |
| Mixed Black Caribbean-White | 1·02 (0·69-1·50) | 1·07 (0·73-1·58) | 1·16 (0·85-1·59) | 1·09 (0·80-1·50) | 0·99 (0·64-1·54) | 1·03 (0·66-1·60) |
| Any other Black background | **0·56 (0·42-0·74)** | **0·56 (0·43-0·75)** | 0·85 (0·67-1·07) | 0·81 (0·64-1·04) | **0·60 (0·43-0·82)** | **0·61 (0·44-0·84)** |
| Bangladeshi | **0·60 (0·44-0·82)** | **0·58 (0·43-0·79)** | 1·14 (0·88-1·47) | 1·07 (0·82-1·38) | **0·67 (0·47-0·95)** | **0·62 (0·44-0·89)** |
| Indian | 0·78 (0·60-1·01) | **0·75 (0·58-0·98)** | **1·24 (1·00-1·54)** | **1·26 (1·01-1·56)** | 1·09 (0·79-1·52) | 1·07 (0·77-1·48) |
| Pakistani | **0·71 (0·58-0·87)** | **0·70 (0·57-0·85)** | 1·12 (0·95-1·33) | 1·09 (0·92-1·29) | 0·91 (0·72-1·15) | 0·89 (0·71-1·13) |
| Chinese | 0·67 (0·36-1·23) | 0·61 (0·33-1·12) | 1·06 (0·63-1·78) | 1·10 (0·65-1·85) | **0·50 (0·26-0·94)** | **0·46 (0·24-0·87)** |
| Mixed Asian-White | **0·50 (0·31-0·81)** | **0·51 (0·51-0·82)** | 1·05 (0·69-1·60) | 0·95 (0·62-1·46) | **0·52 (0·30-0·89)** | **0·50 (0·29-0·87)** |
| Any other Asian background | **0·72 (0·57-0·91)** | **0·71 (0·56-0·90)** | 1·01 (0·83-1·22) | 0·97 (0·80-1·17) | 0·81 (0·62-1·06) | 0·78 (0·60-1·02) |
| Any other mixed background | **0·70 (0·51-0·95)** | **0·70 (0·51-0·95)** | 0·79 (0·61-1·02) | **0·72 (0·56-0·93)** | **0·70 (0·49-0·99)** | **0·68 (0·48-0·97)** |
| Any other ethnic background | **0·66 (0·52-0·84)** | **0·66 (0·52-0·85)** | **0·78 (0·64-0·96)** | **0·76 (0·62-0·94)** | **0·62 (0·47-0·81)** | **0·62 (0·47-0·82)** |
| Unknown or refused | **0·71 (0·59-0·86)** | **0·70 (0·58-0·85)** | 0·91 (0·76-1·09) | 0·87 (0·73-1·04) | **0·77 (0·62-0·95)** | **0·74 (0·60-0·92)** |
| *Gender* | | | | | | |
| Female | 1·28 (1·18-1·39) | **1·27 (1·16-1·38)** | 1·05 (0·98-1·12) | **1·12 (1·04-1·20)** | **1·28 (1·16-1·40)** | **1·28 (1·16-1·41)** |
| Other | 0·57 (0·22-1·50) | 0·56 (0·21-1·49) | 1·32 (0·51- 3·39) | 1·13 (0·43-2·93) | 1·56 (0·41-5·99) | 1·48 (0·39-5·63) |
| Male | 1 | 1 | 1 | 1 | 1 | 1 |
| *Age* | | | | | | |
| <26 | **0·86 (0·78-0·94)** | **0·83 (0·75-0·91)** | **1·34 (1·23-1·45)** | **1·31 (1·21-1·42)** | 1·00 (0·90-1·12) | 0·95 (0·85-1·06) |
| 36-45 | 1·12 (0·99-1·26) | 1·10 (0·98-1·25) | **0·84 (0·76-0·92)** | **0·82 (0·75-0·91)** | 1·10 (0·95-1·26) | 1·08 (0·94-1·24) |
| 46-55 | 0·97 (0·84-1·12) | 0·90 (0·78-1·04) | **0·75 (0·67-0·84)** | **0·72 (0·64-0·81)** | 0·94 (0·79-1·11) | 0·86 (0·73-1·02) |
| 56-66 | 0·98 (0·82-1·18) | 0·88 (0·73-1·06) | **0·73 (0·63-0·84)** | **0·69 (0·59-0·80)** | 0·86 (0·69-1·05) | **0·77 (0·62-0·95)** |
| *Occupational status* | | | | | | |
| In work, education, or training | **1·28 (1·18-1·39)** | **1·31 (1·20-1·43)** | **1·26 (1·17-1·34)** | **1·19 (1·11-1·27)** | **1·48 (1·34-1·63)** | **1·48 (1·34-1·64)** |
| Not in work, education, or training | | | | | | |
| *Caseload size of care coordinators* | 0·99 (0·96-1·01) | 0·99 (0·96-1·02) | **0·97 (0·95-0·99)** | **0·97 (0·95-0·99)** | **0·97 (0·94-1·00)** | **0·97 (0·94-1·00)** |
| *Inequalities strategies* | | | | | | |
| Yes | 1·02 (0·72-1·44) | 1·03 (0·72-1·47) | 0·89 (0·67-1·18) | 0·86 (0·65-1·14) | 0·98 (0·69-1·41) | 0·98 (0·68-1·40) |
| No | 1 | 1 | 1 | 1 | 1 | 1 |
| **Random Part of the Model** | | | | | | |

| | | | | | | |
|---|---|---|---|---|---|---|
| *Between EIP team variance* | | | | | | |
| Year | | | | | | |
| *ICC* | | | | | | |
| Null | **0·41 (0·34-0·47)** | **0·41 (0·34-0·47)** | **0·35 (0·29-0·41)** | **0·35 (0·29-0·41)** | **0·40 (0·34-0·47)** | **0·40 (0·34-0·47)** |
| Unadjusted/adjusted | **0·41 (0·35-0·48)** | **0·42 (0·35-0·49)** | **0·35 (0·30-0·41)** | **0·36 (0·30-0·42)** | **0·41 (0·34-0·48)** | **0·41 (0·34-0·48)** |

OR: Odds ratio; 95%CI: 95% confidence interval. P-values (p<0·05) in bold.

[1]Univariable regression analyses including the outcome and random effects.

[2]Full adjusted model including fixed effects for ethnicity, gender, age, occupational status, caseload size of care coordinators, and inequalities strategies, and random effects for EIP teams and year of data collection.

[3]Caseload size is scaled as the odds associated with one extra patient per care coordinator (i.e., increased caseload size of care coordinators is associated with lower odds of receipt of either psychological intervention).

[4]ICC reported from an unadjusted model including ethnicity as the single covariate.
